# Bismuth subsalicylate profoundly alters gut microbiome and immunity with increased susceptibility to infection

**DOI:** 10.1101/2025.10.01.25337000

**Authors:** Victor I. Band, Phoebe LaPoint, Shira Levy, Lauren Krausfeldt, Ian S. Lacroix, Audrey Chong, Nathan T. Brandes, Benjamin Schwarz, Shreni Mistry, Andrew S. Burns, Mickayla Bacorn, Keona Banks, Rachel Strength, Qing Chen, Hector N. Romero-Soto, Ruhika Prasad, Richard Apps, Liya Wang, Iyadh Douagi, Jessie J. Polanco, Aparna Kotekar, Amrita Mukherjee, Galina Koroleva, Ashley Compean, Brian Sellers, Thomas Langowski, Kevin Rose, Susan Roy, Sivaranjani Namasivayam, P. Juliana Perez-Chaparro, Joanna Chau, Apollo Stacy, Yasmine Belkaid, Suchitra K. Hourigan

## Abstract

Bismuth subsalicylate (BSS) is a commonly used over-the-counter medication for a variety of gastrointestinal symptoms. BSS sequesters gut sulfides, which have been shown in murine studies to be key regulators of gut microbiota. Here, we investigated the impact of BSS on human gut microbiome, immunity and susceptibility to enteric pathogens. We observed a significant shift in microbiome composition after BSS usage, with a profound expansion in bacteria with pathogenic potential including Pseudomonadota. Metabolite composition was greatly altered with increased amino acid levels and decreased short chain fatty acids and secondary bile acids. Notably, there was a collapse of key CD4 T cell subsets in the ileum. Finally, mouse and *ex vivo* human models revealed that BSS treatment increases susceptibility to colonization with the enteric pathogen *Salmonella enterica*. This study underlines the key role of sulfides in human gut microbiome and immunity and warrants further investigation into commonly used sulfide-depleting drugs.

## Main

The gut microbiome is a highly dynamic commensal system that has a strong influence on host functionality and immunity, thereby contributing to host protection against pathogenic infection^1^. A key role of the microbiome is the production of microbial metabolites, such as short chain fatty acids (SCFAs) and secondary bile acids, which have a significant influence on host homeostasis and gut function^2,3^. In addition, the microbiome produces several gaseous metabolites, which have recently been appreciated as an important class of signaling molecules known as ‘gasotransmitters’^4,5^. Among these, hydrogen sulfide (H_2_S) has emerged as a multi-functional metabolite, produced by both host tissue and the microbiome^6^. While systemic levels of free H_2_S exist in nanomolar concentrations, the gut can contain several orders of magnitude higher sulfide concentrations^7^. The role of such a high concentration of this reactive gas in the gut is poorly understood, especially in the context of the function of the microbiome.

The concentration of H_2_S in the gut can vary greatly within individuals due to significant impacts from environmental exposures. These exposures include dietary intake of sulfur amino acids, enriched in animal proteins^8^, as well as pharmaceuticals which alter gut sulfide levels. Bismuth subsalicylate (BSS) is a drug which reacts with free H_2_S in the gut to form bismuth sulfide, trapping and eliminating gut sulfide^9^. BSS is a frequently used, widely available, over-the-counter oral medication for a variety of gastrointestinal (GI) symptoms. It received initial approval by the US Food and Drug Administration (FDA) in 1939 and is now approved for 1) relief of diarrhea, 2) relief of upset stomach, and 3) relief of travelers’ diarrhea^10,11^. It is also used off-label for *Helicobacter pylori* eradication and prevention of traveler’s diarrhea^12–15^. There are typically few reported adverse reactions from the recommended dose. As this medication is sold without a prescription, the true usage is difficult to accurately estimate, but over 20 million units are sold every year in the USA^16^. In mouse models we have previously shown that manipulation of sulfide levels, including via BSS treatment, significantly impacted gut microbiome and immunity ^5,17^. However, how this commonly used, sulfide-depleting drug impacts human gut microbiome and immune function is currently unknown. Here, we aimed to assess the impacts of BSS usage on gut microbiome composition, function and host immunity in healthy human subjects.

## Results

### Participants and study design

To determine how BSS impacts human gut biology, 20 healthy adult subjects were given the maximum recommended dose of BSS orally, 4 doses of 1050 mg daily for 2 days (demographic and clinical data in **Supplementary Table 1**). Fecal and serum samples were collected at baseline (>1 week prior to treatment), directly prior to treatment (day 0) and at days 2, 8, 14 and 28. Additionally, some subjects had optional colonoscopies for retrieval of terminal ileum biopsies at baseline and day 8. In total, 34 subjects were enrolled from September 2023 to October 2024, 13 subjects screen failed prior to BSS administration and one subject withdrew after partial dosing of BSS. There were mild anticipated adverse events related to BSS use in 80% of subjects, with the most common being darkening of stools (**Supplementary Table 2**).

### BSS treatment causes profound, transient changes in microbiome composition

To determine if BSS treatment impacted the composition of the microbiome, fecal samples were analyzed by shotgun metagenomic sequencing. Clear changes were observed in microbiome composition as measured by alpha diversity (**Fig. 1a and Extended Data Fig. 1a-d**) and beta diversity (**Fig. 1b**) after BSS treatment. Alpha diversity was significantly decreased at day 2 (**Extended Data Fig. 1a-d**), with a ∼40% decrease in number of observed species (**Fig. 1a**). Beta diversity was significantly different at day 2 post-BSS treatment, as measured by Bray-Curtis dissimilarity (**Fig. 1b**). At all other time points, on day 8, 14 and 28, we observed no significant differences in either alpha or beta diversity (**Fig. 1a, 1b and Extended Data Fig. 1a-d**), indicating a rapid recovery of microbiome composition after the completion of BSS treatment.

**Figure 1.**
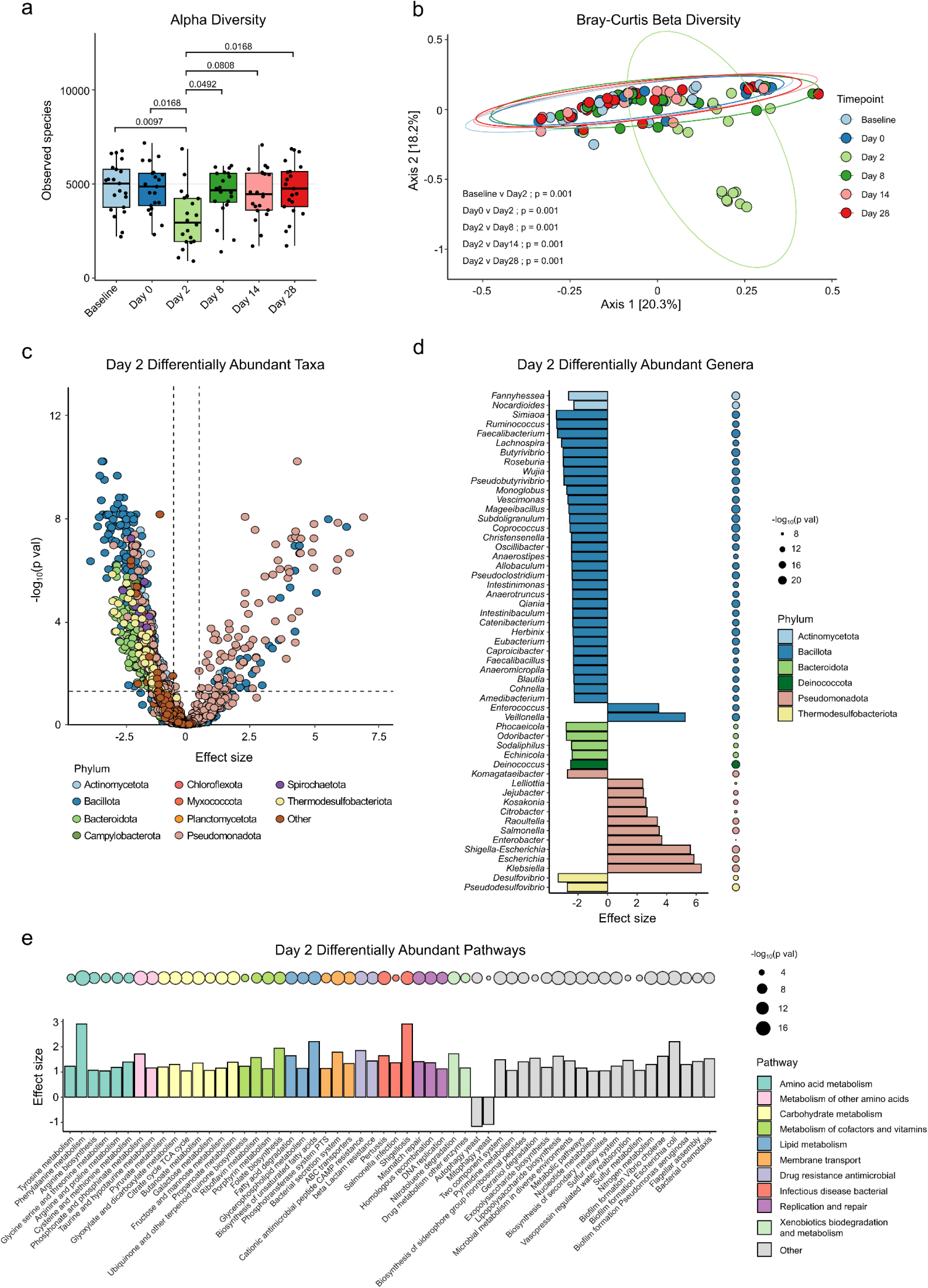
BSS treatment results in profound short-term changes in gut microbiome composition. a, Alpha diversity of fecal microbiome as measured by observed species at each time point. Statistically significant differences in observed species between timepoints are shown with p values adjusted for multiple comparisons from Dunn’s test. **b,** Principal Coordinates Analysis of fecal microbiome beta diversity depicting differences in microbial community composition at each timepoint, as measuredf by the Bray-Curtis dissimilarity metric. Ellipses are drawn around 95% percent of the samples for each timepoint. Statistically significant (p < 0.05) differences between timepoints are displayed from pairwise PERMANOVA. **c,** Differentially abundant species at day 2 compared to day 0. P values were adjusted for multiple comparisons with Benjamini-Hochberg procedure (BH). Dotted lines indicate significance threshold of 0.05 and fold change thresholds of 1 and-1. **d,** Top significant differentially abundant genera. P values were adjusted for multiple comparisons with BH and differences with effect size <-2.25 or > 2.25 are shown. **e,** Top differentially abundant functional pathways in metagenome of fecal microbiome at day 2 compared to day 0. Differences with an effect size of > 1 or <-1 and p value < 0.05 after adjustment for multiple comparisons with BH are shown. Effect size (panels c-e) represents coefficient produced from MaAslin2, representing strength and direction of relationship

At day 2 post-BSS there were 1198 differentially abundant species-level taxa compared to day 0 (**Fig. 1c**), with a significant increase in genera within the Pseudomonadota phylum and a decrease in genera within Bacillota and sulfide-producing Thermodesulfobacteriota phyla (**Fig. 1d, Extended Data Fig. 1e**). Pseudomonadota contains many potential pathogens, and we observed enrichment of *Citrobacter, Salmonella*, *Enterobacter*, *Escherichia* and *Klebsiella* in the gut of BSS-treated subjects (**Fig. 1d**).

*Enterococcus* was also significantly increased, as previously observed in mice (**Extended Data Fig. 2**)^5^. There was a depletion of *Lactobacillus*, key members that promote colonization resistance against pathogens^18,19^ (**Extended Data Fig. 2**). Functionally, there was an expansion of several pathways in amino acid metabolism, carbohydrate metabolism (**Fig. 1e**) and antibiotic resistance pathways and genes (**Extended Data Fig. 3a**). Additionally, we observed depletion of eukaryotic genera (**Extended Data Fig. 3b**) and decreased diversity of the virome which coincided with similar changes in the predicted bacterial host species of these viruses (**Extended Data Fig. 3c-d**).

### Microbiome alteration is associated with significant changes in fecal metabolites post-BSS treatment

The microbiome is a significant source of metabolic activity in the gut, which can have impacts on both local and systemic metabolite availability^20^. To assess how the significant changes in microbiome composition impacted the metabolic landscape, we performed broad targeted metabolomics with chromatography tandem mass spectrometry (LC-MS/MS) assays, focusing on microbially processed and produced metabolites in the gut including the short-chain fatty acids (SCFAs) and bile acids. In agreement with our metagenomics data, we observed significant changes in the gut metabolome at day 2 post-BSS, which quickly subsided at subsequent timepoints (**Fig. 2a**). Among the most significant changes were increases in free amino acid availability, including sulfur-containing amino acids cysteine, cystine and methionine. Nucleic acid metabolites were overall decreased in the gut, though there was an increase in xanthine which is often associated with inflammation (**Fig. 2a**)^21^. SCFAs, key metabolites produced by the microbiome with immune regulatory properties^3^, were significantly depleted in the feces at day 2 post-BSS (**Fig. 2a**). The microbiome also plays a role in modifying bile acids, converting primary bile acids into secondary bile acids^2^. There were large changes in the availability of bile acids (**Fig. 2b**), with an overall decrease in the ratio of secondary to primary bile acids (**Fig. 2c**), indicating a deficiency in the processing of bile acids by the microbiome.

**Figure 2.**
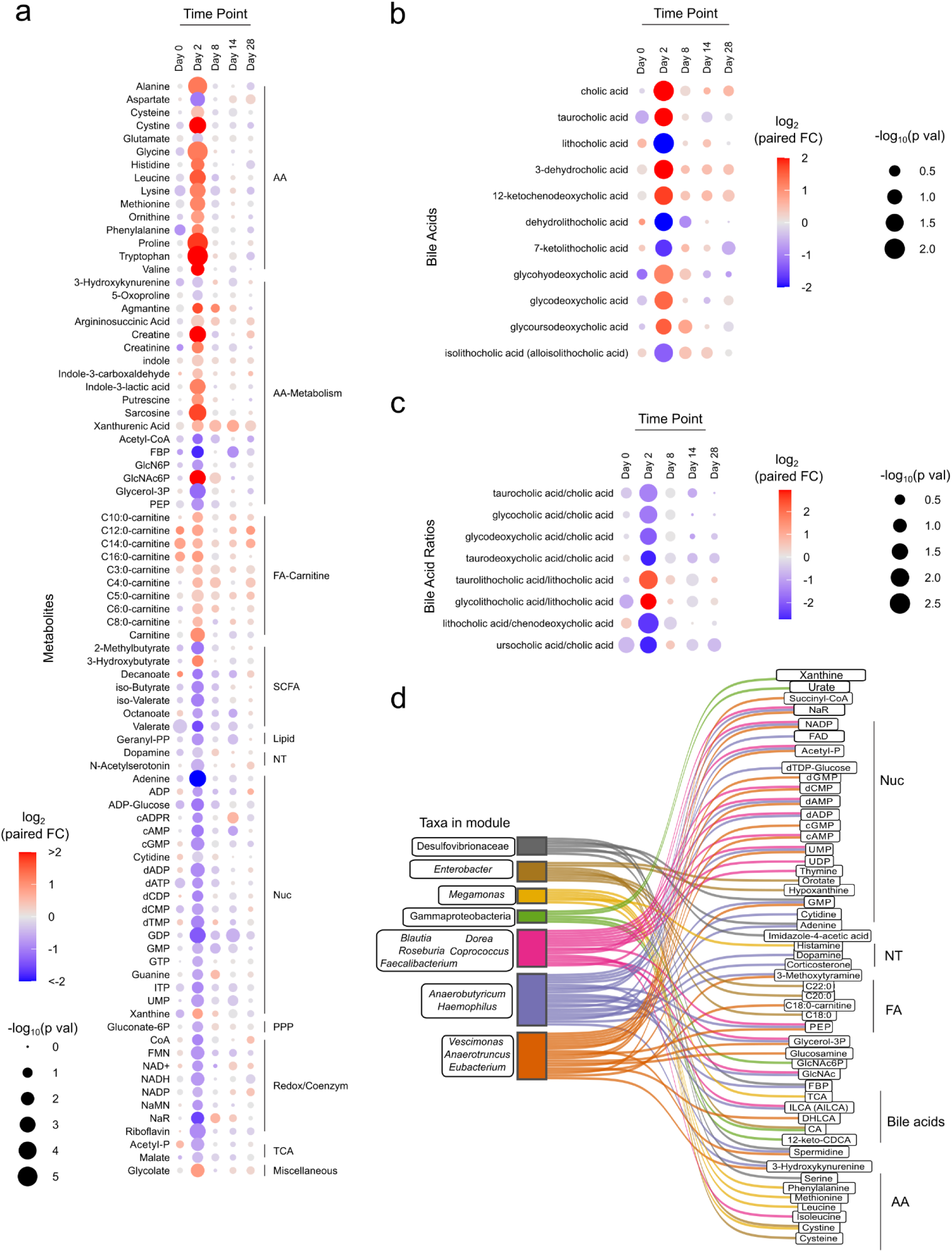
Significant metabolic changes observed after BSS treatment. a, Bubble plot of metabolomics and short chain fatty acids. Fold changes were calculated using paired time point samples, Day 2/Day 0. Bubble color reflected the log2 transformed fold changes. Significant difference between paired time point samples were calculated using the Wilcoxon test. Bubble size reflected the-log10 transformed adjusted p-values. Metabolites included in the plot passed an FDR 10% significance threshold after adjustment using the Benjamini-Hochberg method. Metabolites were arranged by their associated pathway. **b,** Bubble plot of bile acids, and **c,** calculated bile acid ratios. Bubble color reflected paired Day2/Day0 fold changes, and size reflected significance as described in (**a**). Displayed molecules and ratios pass FDR filter of 20%. **d,** Sankey diagram depicting positive correlations between genus level eigengene modules to metabolites, arranged by their associated pathway. Genus eigengene module to metabolite connections displayed in the Sankey diagram had a correlation estimate (Pearson method) greater than or equal to 0.4 with p-value less than 0.05.

To assess which microbes were contributing to these changes in gut metabolite availability, we clustered genera into eigengene modules and assessed associations to each metabolite. Significant contributors included sulfur-cycling Desulfovibrionaceae, which correlated with polyamine spermine, and Gammaproteobacteria, which was associated with increased xanthine and urate abundance, indicating a potential link to inflammation (**Fig. 2d)**. Collectively, these data reveal that BSS induced concomitant changes in the microbiome, microbe-derived immune regulatory metabolites, and metabolites that may indicate inflammation.

### Response to BSS associated with pre-exposure presence of sulfur-cycling taxa

Although we observed significant changes in the microbiome and metabolome of all subjects 2 days post-BSS treatment, it was evident that there was a range of responses among individuals (**Fig. 1b**). While there were some taxa changes seen across all individuals (**Extended Data Fig. 4**) with BSS use, there was a subset of subjects with more pronounced changes as evident by microbiome beta diversity at day 2 (**Fig 1b**), with lasting changes through day 8 and some taxa affected through day 28 (**Extended Data Fig. 5**). We assigned subjects as ‘Low Responders’ (n=12) or ‘High Responders’ (n=8) based on this observation, and in particular the abundance of Pseudomonadota taxa at day 2 (**Fig. 3a**). We observed no association between the magnitude of the microbiome response to any subject metadata (**Supplementary** Figure 1).

**Figure 3.**
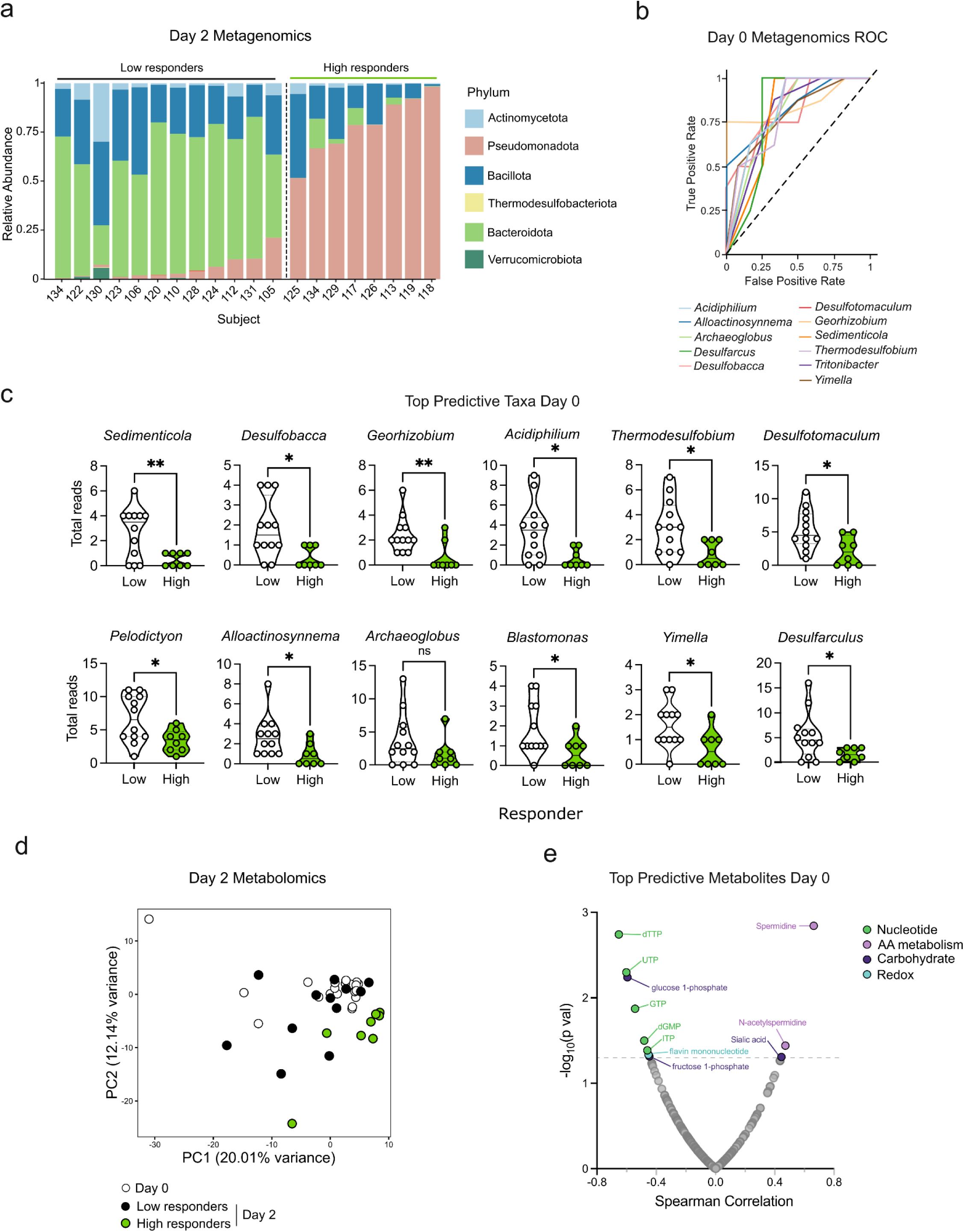
Microbiome composition and fecal metabolites predict magnitude of response to BSS. a, Bar plot depicting relative abundance of top phyla in the microbiome of low and high responders. **b,** ROC curve of top 12 predictive taxa at day 0 for the high responder phenotype. **c,** Top 12 predictive taxa at day 0 for low vs high responders, with sulfur cycling taxa highlighted. **d,** Metabolomics PCA plot at day 0 and day 2 of low and high responders. **e,** U-plot of predictive metabolites at day 0 associated with day 2 Pseudomonadota counts. Correlations were performed using the Spearman method. The dashed grey line indicated a significance threshold of p=0.05. Metabolite point color reflected associated pathways. * p < 0.05, ** p < 0.01, n.s. not significant, unpaired t test.

We then assessed whether the pre-existing microbiome at day 0, prior to BSS treatment, could be associated with the magnitude of response to BSS. Several pre-existing microbial taxa at day 0 were associated with the responder phenotype, as determined by receiver operating curve (ROC) analysis **(Fig. 3b)**. Forty-five total taxa were significantly associated with the responder phenotype (**Supplementary Table 3, Extended Data Fig. 6**), with the majority enriched in low responders compared to high responders. Of the top 12 associated taxa, 8 were known sulfur cyclers, including sulfate reducing microbe *Desulfotomaculum* (**Fig. 3c**). These taxa are known to be high producers of hydrogen sulfide in the human gut and environment^22^. This observation suggests that the presence of high sulfide-producing microbes in the gut may protect against the impact of sulfide depletion on microbiome composition.

Consistent with the microbiome data, the metabolome data also uncovered a subset of subjects with a more pronounced response, closely matching the ‘High responders’ as determined by the microbiome changes (**Fig. 3d, Extended Data Fig. 7**). Reanalysis of the fecal metabolomic data by responder group showed that ‘High Responders’ were responsible for the majority of the observed metabolic changes on day 2 with the exception of increased free amino acids, which were shared in both responder groups (**Extended Data Fig 7**). We next investigated the metabolome to determine if the pre-existing metabolic environment of the gut at day 0 was associated with response to BSS as measured by Pseudomonadota expansion on day 2. Several metabolites were found to be significant, including a positive correlation with regulatory polyamine metabolites spermidine and N-acetylspermidine (**Fig 3e).** Polyamines are a host and microbial produced metabolite family that function as a multifaceted and dynamic regulators of gut homeostasis ^23^. In addition, we observed a negative correlation between day 2 Pseudomonadota and nucleotides GTP and ITP and redox molecule flavin mononucleotide (**Fig. 3e)**. Thus, key differences in the gut microbiome and metabolome were associated with enhanced impact of BSS treatment.

### Reduction in gut T cell subsets after BSS treatment

The gastrointestinal tract is a major site of immune function, with a significant fraction of the body’s immune cells located in this barrier tissue^24^. Previous studies suggest that hydrogen sulfide can act as an important signaling molecule for immune cells^25,26^. Further, we recently observed in mice that sulfide depletion disrupted gut CD4 T cells and mucosal vaccine response^17^. Given these findings, we sought to investigate the potential impact of BSS treatment on gut immune composition. To this end, we sampled gut tissue by pinch biopsy of the ileum during colonoscopy, a site of major gut immune function^27^.

Samples were collected at baseline and 8 days after BSS treatment, with paired samples analyzed from 9 subjects (**Fig. 4a**). Using single cell RNA sequencing on CD45+ cells, we identified 34 distinct clusters including B cell, T cell, plasma cell, and myeloid cell subsets **(Fig. 4b, Extended Data Fig. 8**). We next assessed changes in the frequency of all 34 detected cell subsets and observed that 3 T cell subsets were significantly depleted after BSS treatment: Th1, Th17 and CD8 Trm (**Fig. 4c and 4d**). These subsets also expressed markers compatible with those of tissue resident memory T cells such as *CCL5*, *ITGA1, CD69* and *ITGAE* (**Extended Data Fig. 8**). Due to the significant variation between subjects, we next used pseudobulk analysis on each of the 34 clusters to identify differentially expressed genes (DEGs) among gut immune cells after BSS treatment^28^. This revealed 91 DEGs, including 23 in T cells, 5 in B cells and 11 in myeloid cells (**Fig. 4e, Supplementary Table 4**). Th1 cells were not only reduced in number but also functionality as the remaining Th1 cells had reduced expression of PDK3, a regulator of pyruvate oxidation and the TCA cycle critical for T cell function **(Extended Data Fig. 9)**^29^. In addition, post-treatment Th1 cells had increased MCC expression, a known negative regulator of cell cycle progression **(Extended Data Fig. 9)**^30^. Overall, BSS treatment resulted in a specific depletion of local T cell subsets and altered activation genes implying reduced function from these key immune cells.

**Figure 4.**
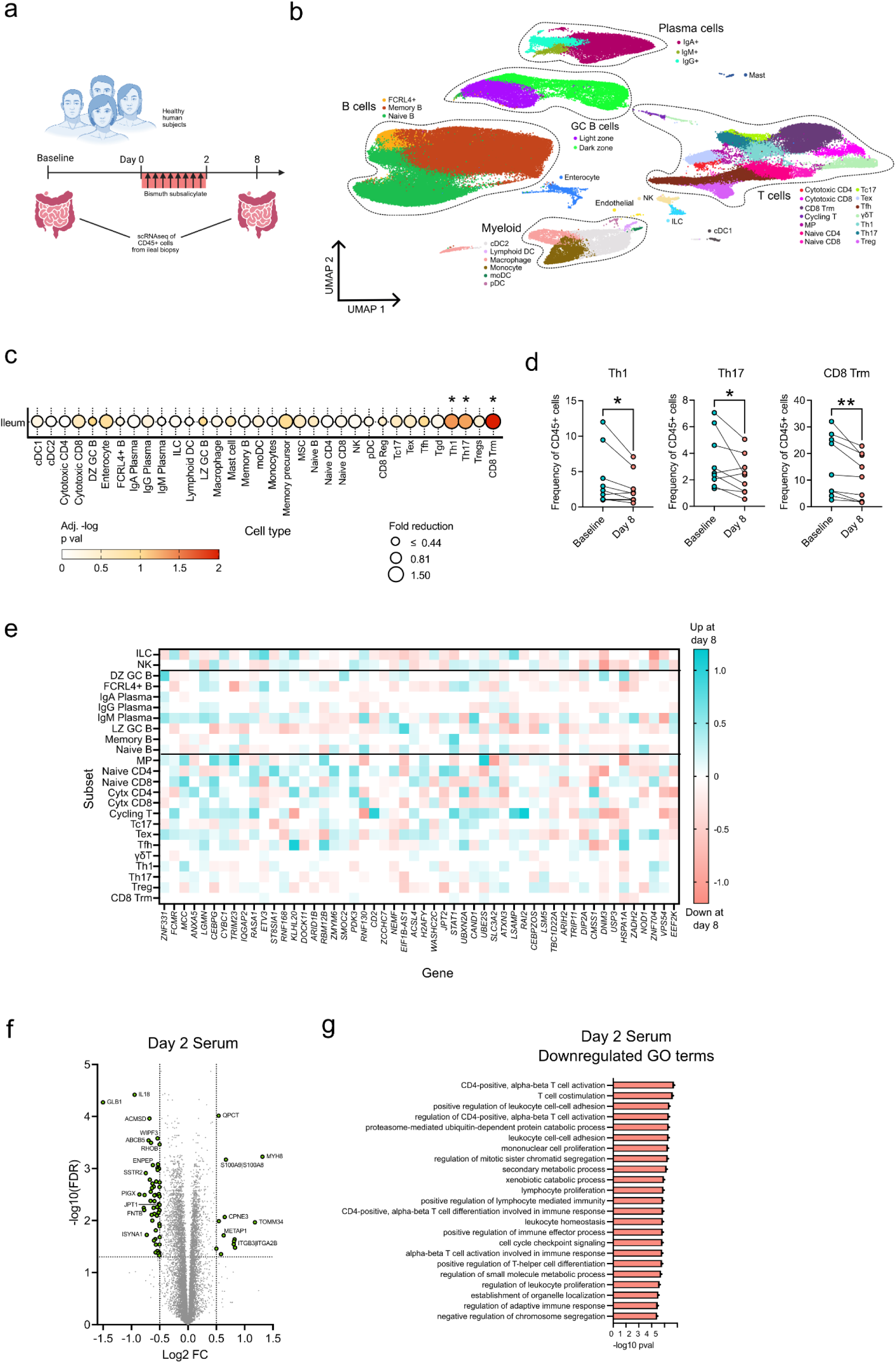
Human subjects treated with BSS demonstrate loss of gut CD4 subsets and systemic proteome alterations. a, Schematic of gut immune sampling in healthy human subjects; subjects underwent colonoscopy with ileal pinch biopsies at baseline and day 8 post-BSS treatment. **b,** UMAP of scRNAseq of ileal biopsies with clusters identified by manual annotation of known immune subsets. **c,** Cell proportions for each identified immune subset in biopsies at day 8 compared to baseline. Size of circles indicates the fold reduction in frequency of day 8 compared to baseline and color indicates p value as measured by paired t test. **d,** Frequency of clusters as proportion of all CD45+ cells, at baseline and day 8. **e,** Heat map of top differentially expressed genes for each cluster among lymphoid cell clusters. **f,** Volcano plot of differentially detected proteins in proteomics of subject serum at day 2 compared to baseline, with color indicating metabolite type. **g,** GO terms enriched for significantly downregulated serum proteins at day 2 compared to baseline.* p < 0.05, ** p < 0.01, paired t test.

Finally, we quantified systemic immune changes using proteomic analysis of the serum at day 2 and day 8 post-BSS treatment compared to baseline. There were significant changes in systemic proteins at day 2, with 13 proteins increased and 55 proteins decreased in serum. These included an increase in the S100A9/S100A8 heterodimeric protein calprotectin, a multifunctional protein that is often a marker of inflammation (**Fig. 4f**)^31^. Gene ontology (GO) term analysis revealed enrichment of several inflammatory pathways, echoing other signatures of inflammation (**Extended Data Fig. 10a**). GO terms downregulated at day 2 included several immune functions, such as CD4 T cell activation and T cell costimulation (**Fig. 4g**). By day 8 post-BSS treatment there were no serum proteins which passed the significance threshold, though gene set enrichment analysis demonstrated a significant increase in several pro-inflammatory pathways including complement activation (**Extended Data Fig. 10 b,c**). We did not observe any enhanced immune changes in the subjects identified as ‘High responders’ in the microbiome data. Overall, BSS usage was associated with a significant impact on gut immune composition and in particular a collapse of Th1 cells, as well as systemic immune changes with evidence for increased inflammation and reduced T cell activation.

### Increased susceptibility to pathogen colonization after BSS treatment

We observed a bloom of potential pathogens in the gut of subjects after BSS treatment and sought to determine whether sulfide depletion with BSS was driving decreased colonization resistance^5^. To this end, we used a mouse model of BSS treatment, where mice were gavaged with BSS at doses scaled down from recommended human dosage **(Fig. 5a)**^32^. When mice were treated with BSS which depleted gut sulfides **(Supplementary** Fig. 2a), we observed significant changes in microbiome composition, similar to the results with human subjects (**Fig. 5b**, **Fig. 1b**). There were significant alterations in beta diversity in the feces and small intestine on day 3, which remained significant beyond day 10 after treatment (**Fig. 5b,c**). Significant depletion of *Lactobacillus* along with increased abundance of *Enterococcus* was observed at day 3, a change also observed in the human microbiome (**Fig.5d, Extended Data Fig. 2**). In mice, *Candidatus* Arthromitus (segmented filamentous bacteria), bacteria with strong immunostimulatory properties, were also significantly depleted.

**Figure 5.**
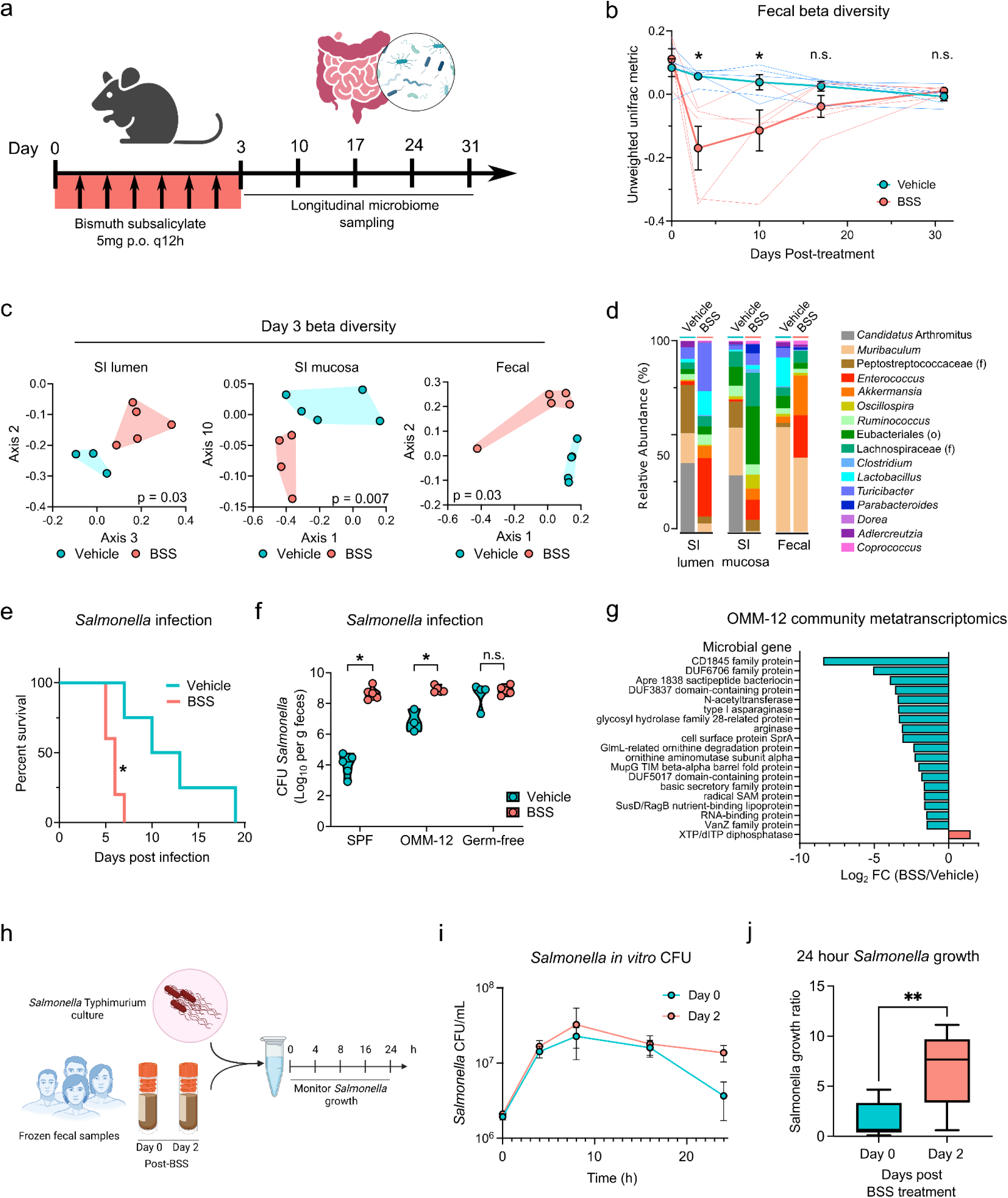
BSS increases susceptibility to Salmonella colonization of the microbiome in mice and humans. a, Schematic of microbiome sampling, mice were treated with BSS then microbiome compartments were sampled weekly for longitudinal analysis. **b,** Longitudinal tracking of fecal microbiome beta diversity in vehicle and BSS treated mice, as measured by unweighted Unifrac distance from 16S sequencing. **c,** Microbiome sampling at day 3 post treatment of small intestinal lumen, mucosa, and feces, as measured by PCA plots of unweighted Unifrac beta diversity. Adjusted p values calculated by PERMANOVA. **d,** Relative abundance of top 16 identified taxa at the genus (or order/family) level in each gut compartment. **e,** Survival of SPF mice infected with *Salmonella* after BSS treatment. **f,** Fecal *Salmonella* CFU in mice after infection following BSS treatment, in SPF, germ free mice, and gnotobiotic mice colonized with the OMM12 defined microbiome. **g,** Fecal metatranscriptomics of OMM12 colonized mice treated with vehicle or BSS at day 4 post treatment. Top significant gene families upregulated (red) or downregulated (teal) in BSS treated mice. **h,** Schematic of *in vitro* infection of human fecal samples with *Salmonella*. **i,j,** *Salmonella* CFU in fecal samples over course of experiment (**i**) and total *Salmonella* growth ratio at 24h growth compared to input at 0h (**j**). * p < 0.05, n.s. not significant, Mann-Whitney U test (b,f), ratio paired t-test (j).

Although the impact of BSS on mice microbiota composition was profound, we did not observe a significant expansion of Gammaproteobacteria, a divergence with our observations in humans. This discrepancy can be explained by the fact that mice used in this study were specific pathogen free with limited colonization with Gammaproteobacteria. To assess the impact of BSS on pathogen colonization, we infected mice orally with gut pathogen *Salmonella* Typhimurium one day after the end of BSS treatment. Mice treated with BSS died significantly faster after infection with *Salmonella* (**Fig. 5e**) and had >4 orders of magnitude higher bacterial burden in the feces at 1 day post-infection (**Fig. 5f**). This increased susceptibility to infection after BSS required the microbiome, as no difference in *Salmonella* infection were observed when germ free mice were treated with BSS (**Fig. 5f**). To further assess the role of the microbiota in these settings, we next colonized germ free mice with the 12-member defined microbiome community OMM12^33^. While OMM12 conferred partial colonization resistance to *Salmonella* infection, BSS treatment reversed the protection afforded by this microbial community (**Fig. 5f**). BSS induced minimal changes in the composition of OMM12 (**Supplementary** Fig. 2b), but was associated with broad alterations to the transcriptome of the microbiome, including loss of bacteriocin production and amino acid metabolism (**Fig. 5g**). Thus, BSS treatment caused both compositional and transcriptional changes in the microbiome of mice, leading to decreased colonization resistance and increased susceptibility to enteric pathogens.

Finally, we sought to determine if the human microbiome post-BSS was more susceptible to colonization with novel gut pathogens. To this end, we collected fecal samples from subjects at day 0 and day 2 of BSS treatment and preserved them in glycerol to maintain viable microbes. We inoculated *Salmonella* Typhimurium into anaerobically thawed and diluted fecal samples and monitored growth over 24 hours (**Fig. 5h**). We selected a subset of ‘Low responder’ subjects which had a minimal amount of Gammaproteobacteria at day 2 post-BSS treatment, as pre-existing Gammaproteobacteria could outcompete the newly introduced *Salmonella*. We monitored the growth and contraction of the *Salmonella* over time in fecal samples from day 0 and day 2 and observed significantly more *Salmonella* CFU at 24 hours in post-BSS feces compared to pre-BSS feces (**Fig. 5i, 5j**). Thus, the fecal microbiome in human samples after BSS treatment was more permissive to *Salmonella* growth and survival. Overall, these data suggest that the profound alterations in the microbiome post-BSS treatment could lead to increased susceptibility to pathogen colonization and enteric infections.

## Discussion

Here, we show the significant impacts of BSS, a commonly used over-the-counter drug, on the composition and function of the human gut microbiome. Notably, potentially pathogenic Pseudomonadota were enriched in these healthy subjects, while *in vitro* and animal experiments suggested increased susceptibility to enteric pathogen *Salmonella*. The significant depletion of *Lactobacillus* is also notable as these taxa play many critical functions in the gut, including controlling inflammation and competing with enteric pathogens^34,35^. In addition, we observed significant changes in local gut and systemic immunity, with a loss of specific immune subsets of Th1, Th17 and CD8 Trm cells in the gut. Systemic immunity also showed evidence for a loss of T cell activation, possibly indicating a widespread impact on these specific T cell subsets. H_2_S has been previously described as a modulator of T cell function^25,26^ which may be an underlying cause of these specific immune changes after BSS. Notably, the T cell subsets we observed to be depleted in this study were also significantly depleted in our prior study of murine gut sulfides^17^. These resident memory subsets play key roles in gut homeostasis, including protection against pathogens and control of inflammation^36^. The compounding of a loss of protective commensals in the gut and the loss of these key immune cells is of particular concern to human gut health and susceptibility to infection after usage of BSS.

BSS has been employed for human use for over 100 years^14^, as a commonly used remedy for nonspecific gastrointestinal symptoms. Despite this, its mechanism of action is poorly understood and has several attested functions that ameliorate GI symptoms. BSS is described to modulate prostaglandins^37^, stimulate reabsorption of fluids and bind to the mucus layer^38^. It is unknown how BSS causes these changes in the gut, though some evidence indicates that it may be driven by the loss of sulfide rather than BSS directly^39^. There is also described antimicrobial activity of BSS^40^, though there is no consensus mechanism of action^41,42^. Intriguingly, the bacteria reported as susceptible to BSS include pathogens which we observed to be enriched in the BSS treated microbiome in our study^43^. Our results indicate that the antimicrobial activity of BSS may be more pronounced in beneficial gut commensals, accompanied by a bloom of potential pathogens. BSS is used off-label to prevent traveler’s diarrhea caused by many of these same pathogens, with evidence for its efficacy in this indication^15,44–46^. However, many of the studies to assess efficacy only look at acute symptoms (diarrhea) and culture of specific pathogens and do not assess potential consequences. Using shotgun metagenomic sequencing to evaluate the whole microbiome, our results show a transient, but profound alteration that permits pathogen colonization, which could potentially have detrimental outcomes in a susceptible host.

In this study, we observed significant microbiome changes across all 20 subjects, however a subset of roughly half of this cohort had a more pronounced response to BSS. The presence of the ‘High responders’ group suggested that there was some factor that potentiated the effects of BSS on the microbiome. We identified the pre-existing microbiome as the main factor that determined the magnitude of the response to BSS, in particular the presence of sulfur cycling taxa which produce high levels of sulfide^22^. We hypothesize that these sulfide producing taxa can complement the loss of sulfide and replenish this lost metabolite after BSS treatment, minimizing the impact on microbiome function. The sulfate-reducing bacteria, among other sulfide producers, have been previously identified as potentially harmful taxa that overproduce H_2_S in the gut and lead to an overly inflamed gut and enhanced disease^47^. However, our study suggests that these taxa may also be beneficial, preventing loss of colonization resistance in the event of sulfide depletion in the gut.

Additionally, we identify several metabolic processes that marked ‘High responders’ including elevated polyamine levels. Polyamines are a host-and microbial-produced metabolite family that function as multifaceted and dynamic regulators of gut homeostasis^23^. Given the multiple sources of input into the polyamine pool it is unclear from these data if observed higher preexisting levels of polyamines in ‘High responders’ are microbial, host, or diet in origin. Interestingly, elevated fecal polyamine levels have been observed in gut pathologies including inflammatory bowel disease and Pseudomonadota species have been annotated as participating in specific conversion of putrescine to spermidine^48^. In addition, this molecule has been reported to enhance colonization of the gut with *Salmonella*^49^.

This study had several limitations. Our sampling of the microbiome in humans only included fecal samples and could not characterize changes in the microbiome in specific parts of the gut. Due to the volatile nature of sulfide and the lack of collection methods to preserve it, we were unable to measure fecal sulfides directly in human samples. Future studies could take advantage of inpatient sampling to test fresh samples or sulfide derivatizing compounds that could preserve sulfide signatures^50^. While we employed a dietary questionnaire to assess changes in diet, the absence of inpatient monitoring limited our ability to control for dietary variations that could impact gut sulfides. We used an over-the-counter formulation of BSS, which contains additives such as flavorings and food dyes which could potentially impact the observed microbiome changes, in addition to the salicylate component of the drug which could be immunomodulatory. However, in our murine experiments we used purified BSS and microbiome changes were consistent with the findings in our human subjects. Healthy subjects between ages 18-50 years were included, and thus we have no data on the impacts of BSS on elderly or pediatric subjects or in those with underlying disease. As with all microbiome studies using read-based classification, which allows for identification of rarer taxa, taxonomic classification of microbes can be suboptimal, especially beyond genus level.

This study highlights that, much like antibiotics, short-term benefits may have unanticipated costs. Our findings that BSS treatment has a significant impact on both gut microbiome and host immunity, even at recommended doses, suggest that we do not fully acknowledge its potential consequences. Further, the fact that these effects were evident in healthy individuals raises the possibility that the unintended consequences of such treatment could be even more dramatic in clinical settings where this drug is typically used, such as treatment of traveler’s diarrhea. This underscores the need for further investigation across broader populations, particularly in individuals with underlying infections, to determine whether these microbiome and immune alterations translate into direct health risks. Beyond resolving acute symptoms, exploring the enduring consequences of any drug that modifies gut physiology and microbial ecology is essential. Given widespread use of BSS and its clear role in shaping microbiome function and pathogen colonization, careful study of its short-and long-term impact is warranted.

## Online Methods

### Subjects, intervention and samples

This study was a single-arm, open-label longitudinal cohort study at the National Institutes of Health (NIH) Clinical Center to evaluate the effect of BSS on the human gut microbiome and host response by studying changes pre-vs. post-drug administration (NCT05930197). Healthy adults were enrolled with written informed consent in the NIH Institutional Review Board (IRB) approved protocol, #001631, “An exploratory study of the effect of bismuth subsalicylate on the gut microbiome and host response in healthy adults”. Subjects were age 18 to 50 years of age, in general good health, without active gastrointestinal disorders and had not taken recent BSS, and without contraindications to taking BSS. Surveys were administered electronically through REDCap. A baseline survey included demographics, clinical data, medical history and dietary information. Follow up surveys included new symptoms following BSS administration and medication use.. BSS dosage timing and compliance were recorded though REDCap.

### Intervention

Subjects were administered BSS suspension at the maximum approved adult dosage (1050mg four times per day) for 2 consecutive days.

### Sample collection

Stool samples were collected at every time point from each subject. Subjects provided samples and survey responses at baseline (between 18 weeks and 1 week before BSS administration), Day 0 (prior to BSS administration, collected up to 7 days earlier), and post-BSS administration at Days 2 (+3 days), 8 (±3 days), 14 (−3/+7 days), and 28 (±7 days). Baseline and Day 0 samples were collected to show stability of microbiome prior to BSS treatment.

Subjects provided a whole stool sample, as well as samples aliquoted in an OMNIgene-gut tube (DNA Genotek In) for DNA preservation and OMNImet-gut tube (DNA Genotek In) for metabolite preservation ^51^. If not provided during the in-person visit, stool samples were collected at home and received at the in-person visit or mailed overnight with ice packs in an insulated shipping container. Once received, whole stool samples were aliquoted into tubes with glycerol (for bacterial preservation), and whole stool without preservatives. Optional blood samples were collected at all time points including a serum aliquot. A subset of the cohort chose to participate in optional colonoscopies at the baseline and day 8 visits. The baseline colonoscopy was performed at least 22 days prior to BSS administration to allow recovery of the microbiome after bowel preparation and adequate subject recovery time between colonoscopies. During colonoscopy, intestinal biopsies were collected from the terminal ileum using cold forceps. Biopsies were collected in media consisting of RPMI1640, GlutaMAX supplement, 10% FBS, penicillin/streptomycin/Fungizone, and 50 μM 2-mercaptoethanol and then transferred into cryovials containing 500ul of Gibco Freezing Media with 10 μM Y-27632. All samples were stored at-80°C after processing until analysis.

### Inclusion Criteria

An individual must meet all the following criteria to be eligible for this study:

1. Aged 18 to 50 years.
2. In generally good health.
3. Able to provide informed consent.
4. Willing to allow samples and data to be stored and shared for future research.
5. Participants who can become pregnant must agree to use one effective method of contraception when engaging in sexual activities that can result in pregnancy, beginning at the signing of the informed consent form (as early as week −18) until the final study visit. Acceptable methods of contraception include the following:

a. External or internal condom with spermicide.
b. Diaphragm or cervical cap with a spermicide.
c. Hormonal contraception.
d. Intrauterine device.

### Exclusion Criteria

An individual who meets any of the following criteria will be excluded from participation in this study:

1. Use of systemic antibiotics in the last 3 months.
2. BSS use in the last 3 months.
3. Pregnant or breastfeeding.
4. Allergy to BSS.
5. Allergy to other salicylates (including aspirin).
6. Current use of other salicylates (including aspirin).
7. Current use of anticoagulant medications.
8. History of or active GI ulcers.
9. History of or active bleeding disorder.
10. Bloody stool within the last 3 months.
11. Diarrhea within the last 2 weeks (defined as three or more loose or liquid stools per day).
12. Current use of medications that may have a drug interaction with BSS
13. Not proficient in written English.
14. Currently participating in another clinical trial that may affect current study procedures, per investigator’s discretion.
15. Any condition that, in the opinion of the study team, contraindicates participation in this study.

## Microbiome Sequencing

### DNA Extraction

DNA was extracted from human and murine fecal samples in two stages. First, approximately 50 mg of fecal material and 650 mL MBL lysis buffer from the Qiagen PowerMicrobiome DNA/RNA EP Kit were added to Lysis Matrix E (LME) tubes (MP Biomedicals). LME tubes were transferred to a Precelleys 24 Tissue Homogenizer (Bertin Technologies) and fecal samples were homogenized, centrifuged, with the resultant supernatant transferred to a deep-well 96-well plate. The second stage consisted of DNA isolation from the above supernatant using the Qiagen MagAttract PowerMicrobiome DNA/RNA EP Kit on an Eppendorf automated liquid handling system as detailed by the manufacturer.

### Metagenomic Sequencing

Total DNADNA content of the microbiome was assessed through shotgun metagenomic sequencing. Metagenomic libraries were constructed from 100 ng of DNA as starting material using the Illumina DNA Prep kit. Illumina DNA/RNA UD Indexes were used to add sample-specific sequencing indices to both ends of the libraries. An Agilent 4200 TapeStation system with High Sensitivity D5000 ScreenTape (Agilent Technologies, Inc) was used to verify quality and assess final library size. Final pools were diluted to 750 pM and sequenced on a NextSeq1000 sequencer using a paired-end (150×150) NextSeq 1000/2000 P2 (300 cycles) kit (Illumina).

### Data processing

Raw sequence data was evaluated for quality and adapters were trimmed using Nephele’s Short read quality check pipeline. Trimmed data was then processed through Nephele’s (nephele.niaid.nih.gov) WGSA2 pipeline ^52^. Briefly, contaminating host reads were removed with kraken2 (v2.1.3) against a *Homo sapiens* and *Mus musculus* database. Remaining reads were taxonomically classified against Standard Reference Genome database (as of March 2023) to produce an abundance matrix of species counts. Reads were also assembled and then mapped back to assemblies. Genes were called and annotated with the Kyoto Encyclopedia of Genes and Genomes (KEGG) database to assign KEGG Orthology (KO) IDs. Copies per million (CPM, equivalent to transcript per million [TPM]) were calculated for each gene and summed for each KO ID to produce an abundance matrix of annotated genes. Functional inference was performed with MinPath (v1.6), and the CPM of genes identified for each pathway were averaged to produce an abundance matrix of functional pathways. Antimicrobial resistance (AMR) genes were identified with AMRFinder (v3.12.8) and CPM of each AMR gene were summed per accession number to produce an abundance matrix of AMR genes using a custom python script.

Assemblies generated from WGSA2 were processed through Nephele’s DiscoVir pipeline for virome analysis^52^. Briefly, viral genomes were identified with default parameters for geNoMad filter and a minimum sequence length of 5000 bp. All viral genomes were clustered at 95% identity and 85% alignment to produce viral OTUs (vOTUs, representing viral species).

CPM of each vOTU were generated. Predicted hosts of vOTUs were identified with minimum confidence score of 90, and vOTUs per highest taxonomic classification of host (i.e. genus, family) were summed to produce an abundance matrix of viruses by their predicted host.

### Statistical analysis

Raw counts of bacterial and archaeal species were filtered for prevalence (10%) and abundance (count of 10) and then rarefied at 5,900,000 reads to normalize for differences in library size. Potential variables to consider as confounders and effectors were evaluated using Permutational Multivariate Analysis of Variance (PERMANOVA) for beta diversity and Kruskal-Wallis or Dunn’s test for alpha diversity. Covariates with days post-BSS dosing were also evaluated with a Chi-squared test. Each variable was included in the model with days post-BSS,age, sex, race, ethnicity, BMI, and if a colonoscopy was performed prior or post-BSS dosing, which were not confounders or effectors of the main treatment.

Differences in Observed species, Evenness, Shannon, Inverse Simpson, and Chao1 for alpha diversity over time due to BSS dosing were evaluated for statistical significance after multiple comparisons with Dunn’s test. Differences in beta diversity were evaluated with pairwise PERMANOVA. Differentially abundant taxa at each day post-BSS dosing compared to day 0 were identified using MaAsLin2 with linear model after log transformation. Taxa were considered significantly different from day 0 with a coefficient of > 1 or <-1 and a p value of 0.05 after adjustment for multiple comparisons (q value), unless otherwise noted. When analyzing taxa at different taxonomic levels, species counts were summed within the group to which it belongs, and the same statistical analyses were performed.

Eukaryotic species counts were filtered for prevalence (10%) and abundance (count of 10) and normalized to relative abundance due to the low number of totals read annotated as eukaryotic. Alpha and beta diversity of eukaryotic species were assessed in the same way as bacteria and archaea, and differentially abundant genera were identified with MaAsLin2 using the same cut offs.

Functional pathways, AMR genes, and viruses classified by their predicted bacterial hosts were filtered for prevalence (10%) and abundance (CPM of 3). They were assessed for compositional differences using pairwise PERMANOVA and differential abundance was performed each day post-BSS dosing compared to day 0 with MaAsLin2 with a coefficient of > 1 or <-1 and a p value of 0.05 after adjustment for multiple comparisons. When analyzing AMR genes at different classification levels (*i.e.* gene IDs), CPMs for all accession numbers for that classification were summed.

All statistics were performed in R using the packages vegan, phyloseq, pairwiseAdonis, and MaAslin2A ^53–55^.

### Evaluation of high and low response to BSS

High and low responders were identified based on distinct clustering of bacteria and archaea in the PCoA. Differential abundance using MaAsLin2 were performed separately on both high and low responders to BSS to identify taxa that changed in both groups. P values after adjustment for multiple comparisons of < 0.05 and < 0.2 was considered significant for high and low responders, respectively, with a coefficient of > 1.5 or <-1.5.

Receiver Operating Characteristic analysis was performed on day 0 counts of genera using the R package pROC to predict a high or low response at day 2 after BSS dosing. Genera with an area under the curve (AUC) of 0.70 and upper and lower confidence intervals of 0.50 were evaluated with a t test for differences in fold change abundances. Those that were statistically significant p < 0.05 were considered predictive of the response at day 2.

### Correlation networks

Counts of genera were clustered using weighted gene co-expression network analysis (WGCNA) in the R package WGCNA to create representative “module eigengenes” with associated abundances to correlate with metabolome. WGCNA was performed using a power of 7, deepSplit of 2, and minimum module size of 10. These module eigengenes were defined by the most abundant genus or genera within that module that significantly correlated to the module eigengene abundance. The same was done with gene abundances to create module eigengenes, using a power of 20, deepSplit of 3, and minimum module size of 20.

## Metabolomics

### Stool Sample Preparation

For all LC-MS/MS methods LC-MS/MS grade solvents were used. All samples were immersed in 0.4 mL of ice-cold methanol. To each sample 0.4 mL of water and 0.4 mL of chloroform were added. Samples were shaken for 30 minutes under refrigeration and centrifuged at 16000 x g for 20 min. 400 µL each of the top (aqueous) layer and bottom (organic) were collected separately. A subaliquot of the aqueous layer was taken for O-benzylhydroxylamine derivatization of carboxylic acids and short chain fatty acid analysis. The remaining aqueous layer was diluted 5x in 50 % methanol in water for LCMS analysis of central polar metabolites. The organic layer was dried down under vacuum and resuspended in an equivalent volume of 5 µg/mL butylated hydroxytoluene in 6:1 isopropanol:methanol for bile acid analysis.

### Short Chain Fatty Acid Derivatization

Samples were derivatized with O-benzylhydroxylamine (O-BHA) according to previously established protocols. ^56,57^ Reaction buffer was prepared fresh consisting of 1M pyridine and 0.5 M hydrochloric acid in water. A 35 µL aliquot of the aqueous extract was taken and to the sample was added 10 µL of 1M O-BHA in reaction buffer and 10 µL of 1M 1-Ethyl-3-(3-dimethylaminopropyl) carbodiimide in reaction buffer. Samples were shaken at room temperature for 2 hrs. Each sample was quenched with 50 µL of 0.1 % formic acid for 10 min. Derivatized carboxylic acid compounds were extracted with the addition of 400 µL ethyl acetate. Samples were centrifuged at 16000 x g for 5 min at 4 °C to induce layering and the upper (organic) layer was collected. The extract was dried under vacuum and each sample was resuspended in 300 µL of water for LCMS injection.

### Liquid Chromatography Mass Spectrometry (LC-MS/MS)

Tributylamine and all synthetic molecular references were purchased from Millipore Sigma. LCMS grade water, methanol, isopropanol and acetic acid were purchased through Fisher Scientific.

Aqueous metabolites were analyzed using a combination of two analytical methods with opposing ionization polarities ^58,59^. Both methodologies utilized a LD40 XR UHPLC (Shimadzu Co.) system for separation and a 6500+ QTrap mass spectrometer (AB Sciex Pte. Ltd.) for detection. Negative mode samples were separated on a Waters Atlantis T3 column (100Å, 3 µm, 3 mm X 100 mm) and eluted using a binary gradient from 5 mM tributylamine, 5 mM acetic acid in 2% isopropanol, 5% methanol, 93% water (v/v) to 100% isopropanol over 5 minutes. Two distinct MRM pairs in negative mode were used for each metabolite. Positive mode method samples were separated across a Phenomenex Kinetex F5 column (100 Å, 2.6 µm, 100 x 2.1 mm) and eluted with a gradient from 0.1 % formic acid in water to 0.1 % formic acid in acetonitrile over 5 minutes.

Derivatized short chain fatty acid samples were analyzed using a LD40 XR UHPLC (Shimadzu Co.) system for separation and a 6500+ QTrap mass spectrometer (AB Sciex Pte. Ltd.) for detection. Samples were separated with a Waters™ Atlantis dC18 column (100Å, 3 µm, 3 mm X 100 mm) using a 6 min gradient from 5-80 % B with buffer A consisting of 0.1 % formic acid in water and B consisting of 0.1 % formic acid in methanol. Short chain fatty acids and central metabolic carboxylic acids were detected using positive mode MRMs from previously established methods and identity was confirmed by comparison to derivatized standards.^56,57^

Bile acid samples were analyzed by injecting a fraction of the organic layer from the initial metabolite extraction was injected on a LD40 X3 UHPLC (Shimadzu Co.) and a 7500 QTrap mass spectrometer (AB Sciex Pte. Ltd.) was used for detection.

Peaks were resolved on a Phenomenex Kinetex Polar C18 (100Å, 2.6 µm, 3 mm X 100 mm) using a binary gradient of A: 0.01 % acetic acid in water and B: 0.01 % acetic acid in methanol. A 20 min gradient from 40-100 % B was utilized for separation. Samples were detected in negative MRM mode using previous validated MRMs ^60^. Internal bile acid standard signals (Bile Acid SPLASH, Avanti Polar Lipid) were used to confirm signal identities and retention times.

### Metabolome Data Processing and Bioinformatics

All signals were integrated using SciexOS 3.1 (AB Sciex Pte. Ltd.). Signals with greater than 50% missing values for a specific tissue set were discarded and remaining missing values were replaced with the lowest registered signal value. Where appropriate, signals with a QC coefficient of variance greater than 30 % were discarded. Metabolites with multiple MRMs were quantified with the higher signal to noise MRM. Filtered datasets of the negative mode aqueous metabolites were total sum normalized after initial filtering. The SCFA dataset and the positive mode aqueous metabolomics dataset were scaled and combined with the negative mode aqueous metabolite dataset using common signal for malate and tyrosine respectively. All statistical analyses were performed in R (version 4.5.1). The wilcox_test() function from the rstatix package was used for paired time point comparisons. The rstatix adjust_p_value() function with the Benjamini-Hochberg method was applied where indicated for correction across multiple comparisons. A false discovery rate (FDR) threshold of 10% was applied to these data where indicated. Correlation between metagenomic genus eigengene modules and metabolites were performed using the rcorr() function, with the pearson method, from the Hmisc package. Resulting correlations were filtered to an estimate greater than or equal to 0.4 with a p value less than 0.05. The Sankey diagram was generated using the ggsankey package. Linear regression was performed using the lm() function from the stats package.

## Ileal biopsy analysis

### Single cell preparations from biopsies

All biopsies were processed in parallel and were thawed and immediately washed 2 times in basal organoid media consisting of advanced DMEM/F12 with nonessential amino acids and sodium pyruvate (Thermo Fisher Scientific), 10 mM HEPES (Thermo Fisher Scientific), 2 mM Glutamax (Thermo Fisher Scientific), 100 μg/mL Normocin (Invivogen), 100 U/ml penicillin (Thermo Fisher Scientific), 100 μg/mL streptomycin (Thermo Fisher Scientific), and 1 mM N-acetylcysteine (Thermo Fisher Scientific), supplemented immediately before use with 10 μM Y-27632 (Stemcell Technologies). This represents minor modifications from a previously described basal organoid media ^61^. Samples were then washed in PBS before incubation in enzymatic digestion media at 37C for 30 mins with continuous agitation. Digestion media comprised RPMI1640 (Cytiva), 10 mM HEPES, 100 U/ml penicillin, 100 μg/mL streptomycin all as above, 2% FBS (Thermo Fisher Scientific), and 50 μg/mL gentamicin (Thermo Fisher Scientific), supplemented immediately before use with 100 μg/mL Liberase TM (Roche) and 100 μg/mL DNase I (Roche). This represents minor modifications from a previously described enzymic digestion mix for biopsy samples ^62^. Digestion was stopped with 10mM EDTA and then aspiration with a pipette tip used to disrupt tissue before pressing through a 40μM cell strainer and collection in basal organoid media.

### Pooling strategy for post-digest experimental handling

Samples were combined into 2 pools for post-digest processing. Each pool comprised 1 sample from each of the individuals, enabling subject demultiplexing based on genotype. Pre and post treatment samples from an individual were separated between pools, with scrambling so that pool did not confound timepoint comparisons across all individuals in the same way. A single ileum biopsy from an excluded subject was digested in parallel to the study samples, as well as thawing of cryopreserved PBMC from an additional healthy donor, for inclusion equally in both pools as a control for batch effects between pools. Sample yields enabled a mean of 378k (range 244k - 410k) live cells based on AOPI count from each of the study samples to be used for pool construction, in addition to 250k of the ileum and 50k of the PBMC spike samples.

### Cytometry enrichment of live leukocytes

Post-digest single cell preparations showed mean viability of 50% (range 39% - 62%) for the study samples. In parallel the 2 pools were resuspended in PBS with 2% FBS (FACS buffer) and incubated with True-Stain Monocyte Blocker for 5 mins on ice (Biolegend), before staining for 30mins with CD45-PE (BioLegend). Samples were then washed, resuspended in FACS buffer, and 5 mins prior to sorting DAPI was added (Thermo Fisher). Sorting of CD45+ DAPI-cells was conducted using a SONY MA900. Cells were collected in 10% FBS and post-sort AOPI counts showed viabilities of 90% and 88% for the 2 pools.

### Single-cell capture and sequencing

192k cells from pool 1 and 149k cells from pool 2 were loaded across a total of 8 lanes with reverse transcription mix on a Chromium chip and partitioned into single cells in Gel Beads-in-emulsion (GEMs) using the 10x Genomics Chromium GEM-X Single Cell 3’ Kit v4 as per manufacturer’s instructions (10x Genomics). Gene expression libraries were prepared as per the 10x Genomics protocol (User Guide CG000731, Rev B), with all amplification steps performed in an Applied Biosystems Ins Veriti 96-well thermal cycler. Quality and quantity of the libraries were assessed using TapeStation (Agilent) and a Qubit fluorometer (ThermoFisher). Libraries were pooled at a concentration of 20 nM and sequenced at the Center for Cancer Research sequencing facility (NovaSeq Xplus, Illumina) using 10B 100 cycle flow cell with sequencing depth calculated at 50,000 read pairs/cell.

### Bulk RNA-seq for subject demultiplexing

For each subject bulk RNA-seq was generated for subject demultiplexing of the single cell data. RNA was extracted from PBMC using RNeasy Plus Micro (Qiagen) according to manufacturer instructions. Libraries were generated using NEB Ultra II (NEB), with QC using TapeStation (Agilent) and a Qubit fluorometer (ThermoFisher). Libraries were pooled at a concentration of 2 nM and sequenced at 1 nM (NextSeq 2000, Illumina) using 150bp paired end reads with a sequencing depth of 100M reads/sample.

### Analysis

Eleven subjects underwent colonoscopy and of these, 9 subjects had paired ileal biopsies available at baseline and days 6-11. Each pool of samples was demultiplexed using demuxalot (https://github.com/arogozhnikov/demuxalot) and singlets were selected if the refined posterior probability was over 0.99. For quality controls, cells with less than 200 detected genes, greater than 15% mitochondrial reads, or gene counts greater than 10,000 were removed. Cell clustering was performed by applying the FindNeighbors function on PCA reduction with dimension 1:30, followed by Leiden clustering on the resulting SNN graph using Seurat’s FindClusters algorithm, with a resolution parameter of 0.8. Clusters were identified usingcelldex::HumanPrimaryCellAtlasData for automated annotation via SingleR. Cells were then manually annotated by considering automated annotation and gene expression data (Extended Data Fig. 8) using Loupe Browser (10X Genomics). Differential gene expression was performed using pseudobulking genes by average expression for each identified cluster, timepoint and subject. Differential genes were calculated with DESeq2 using MAST ^63,64^ for each cluster, using subject id as a latent variable to correct for patient-to-patient variability, and reporting genes having an adjusted p value <0.1. All analysis was performed in R using the Seurat package ^65^.

### Proteomic analysis of serum

Proteomic profiles were characterized in human EDTA-plasma samples using the SomaScan Assay v5.0 (SomaLogic Inc., Boulder, CO). This measures ∼11,000 proteins across a wide range of biological processes including cytokines, hormones, growth factors, receptors, kinases, proteases, protease inhibitors and structural proteins. The SomaScan Assay was performed on a Tecan Fluent 780 high throughput automation system according to manufacturer’s instructions at the NIH Center for Human Immunology. Assay data were normalized using SomaLogic’s standard procedure including adaptive normalization by maximum likelihood^66^. For each analyte, paired t-tests were performed between baseline (day 0) and post-treatment (day 2/8) samples with Benjamini–Hochberg correction applied to control the false discovery rate. Log₂ fold changes were calculated to rank analytes. For day 2, significantly up-and down-regulated proteins (p < 0.05) were subjected to Gene Ontology enrichment analysis using clusterProfiler (v4.14.4). For the day 8 time point, genes were ranked by signed p-value (sign(log_2_ fold change) ×-log_10_ (p-value)) and analyzed by Gene Set Enrichment Analysis of Biological Process terms using clusterProfiler (v4.14.4).

## Animal experiments

### Mice and bismuth treatment model

All wild-type mice used were C57BL6/J female and male mice aged 8-14 weeks at the start of the experiment, and were sex and age matched. All mice were obtained from the Taconic-NIAID exchange program. For all experiments, littermate control mice were randomized into cages for 1 week before beginning experimental manipulations to correct for cage effect. Specific pathogen free (SPF) mice were bred and maintained under SPF conditions at an American Association for the Accreditation of Laboratory Animal Care (AAALAC)–accredited animal facility at NIAID and housed in accordance with the procedures outlined in the Guide for the Care and Use of Laboratory Animals. Germ-free and gnotobiotic colonized mice were housed in the NIAID Gnotobiotic Animal Facility under gnotobiotic conditions, and were all bred in the facility. All experiments performed under an approved animal study proposal (LHIM2E), approved by the NIAID IACUC.

Mice were treated with BSS by oral gavage or via drinking water. SPF mice treated by gavage were given 5mg BSS in H**_2_**O orally by gavage, every 12 hours for 72 hours, for a total of 6 doses. Germ-free and gnotobiotic mice were given autoclaved drinking water containing 7mg/mL BSS for 72 hours. Precipitated BSS was resolubilized by shaking every 24 hours. Vehicle treated mice were given H_2_O at identical doses and times.

### 16S rDNA sequencing

Samples were collected fresh from mice and kept at-20°C until sequencing. Fecal samples were collected from live mice in sterile collection cups. Small intestine luminal content was collected at necropsy by removing tissue and shaking vigorously in PBS to dissociate luminal bacteria. Mucosal associated bacteria were then isolated by scraping this tissue with forceps to remove mucus layer and shaken vigorously in PBS. Fractions were then spun down at 5000xg for 10 min to pellet bacteria. Fecal DNA was purified using the MagAttract PowerMicrobiome DNA/RNA kit (Qiagen). Amplification of the V4 hypervariable region of the bacterial 16*S* rRNA gene was performed using the 515f and 806r primers (515F: 5′ - GTGYCAGCMGCCGCGGTAAY-3′; 806R: 5′-GGACTACNVGGGTWTCTAATN-3′), with Illumina sequencing adapter and unique barcodes integrated in each primer. Amplicons were quantified using Kapa Library Quantification Complete Kit (ROX Low) (Kapa Biosystems) and pooled at equimolar concentrations before being sequenced on the Illumina MiSeq.

Illumina DNA/RNA UD Indexes were used to add sample-specific sequencing indices to both ends of the libraries. An Agilent 4200 TapeStation system with High Sensitivity D5000 ScreenTape (Agilent Technologies, Inc) was used to verify quality and assess final library size. Final pools were diluted to 750 pM and sequenced on a NextSeq10001 sequencer using a paired-end (100×100) NextSeq 1000/2000 P2 (200 cycles) kit (Illumina, Inc). 16S sequencing data was analyzed with QIIME2^67^. using the DADA2 denoising algorithm to generate ASVs and the Greengenes 13.8 database for taxonomic classification.

### Salmonella infection

Mice were treated with BSS or vehicle as described above for 3 days. Mice were then fasted overnight and infected 24 hours after final dose of BSS. *Salmonella enterica* serovar Typhimurium strain IR715/NalR was grown overnight from 2-3 colonies in Luria Broth, and 1×10^7^ CFU was administered to mice orally by gavage in 100uL of PBS. Mice were monitored for illness, survival, weight loss and fecal CFU. CFU was determined by homogenizing fecal pellets in sterile PBS and plating on selective LB agar plates containing 50μg/mL nalidixic acid.

### Metatranscriptomics

Fecal pellets from gnotobiotic OMM12 colonized mice were collected at day 3 after BSS or vehicle treatment and stored in RNAlater ICE (Thermofisher) at-80 °C. RNA was isolated from 2 fecal pellets using RNEasy PowerMicrobiome kit (QIAGEN). Ribosomal RNA was depleted with the CORALL RiboCop rRNA Depletion kit (Lexogen) and libraries prepared with CORALL RNA-Seq V2 kit (Lexogen). Libraries were pooled to 650nM and sequenced on a NextSeq 2000 sequencer (Illumina) using 150 bp paired end NextSeq 1000/2000 P2 kit (Illumina). Data was analyzed using the SAMSA2 pipeline ^69^.

### Human *in vitro* fecal infection

Approximately 150mg of fecal samples from patients at day 0 and day 2 were aliquoted into tubes containing 20% glycerol in PBS and stored at-80 °C to preserve live microbes. To conduct the *in vitro* infection, samples were spun down at 10,000xg and approximately 50mg of fecal sample was combined with 2uL PBS per mg of feces. *Salmonella enterica* serovar Typhimurium strain IR715/NalR was grown overnight from single colony in Luria Broth, and 2.5 x 10^6^ CFU/mL were added to fecal solution in closed microcentrifuge tubes. Tubes were incubated at 37 °C and sampled at 0, 2, 4, 8 and 24 hours.

Salmonella CFU was quantified by growth on selective LB agar plates containing 50μg/mL nalidixic acid. All steps of the experiment were conducted in an anaerobic chamber to preserve oxygen sensitive microbes, though initial fecal collection occurred in standard oxygen conditions.

## Data availability

All data generated in this study will be deposited into publicly available databases upon publication.

## Author Information

Victor I. Band, Phoebe LaPoint, and Shira Levy contributed equally to this paper. Suchitra K. Hourigan and Yasmine Belkaid also contributed equally to this paper.

## Contributions

S.K.H., Y.B. and V.I.B. conceived and designed the project. S.L. and S.R. performed project administration. V.I.B, L.K., A.C., N.T.B., B.S., I.S.L., L.W., K.R performed data analysis. P.L., M.B., K.B., R.S., Q.C., H.N.R.S., R.P., S.L. and S.N. performed investigation and data curation. S.M. and A.B. performed sequencing experiments. R.A., J.J.P., A.K., A.M., G.K., A.C., B.S. and T.L., K.R., R.A. (study design) and I.D. (supervision) generated immunologic data. V.I.B., P.J.P.C., J.C. and A.S. performed murine and *in vitro* experiments. S.K.H. and Y.B. obtained funding for the project and supervised the project. V.I.B., P.L., and S.K.H. wrote the original draft. All authors reviewed, edited and approved the final manuscript.

## Corresponding authors

Suchitra K. Hourigan. Laboratory of Host Immunity and Microbiome, National Institute of Allergy and Infectious Diseases, National Institutes of Health, Building 50, Rm 5511, 50 South Dr, Bethesda, Maryland, 20892.

Yasmine Belkaid. Unité Metaorganisme, Immunology Department, Pasteur Institute, Paris, France.

## Ethics approval and consent to participate

The study was approved by an Institutional Review Board (IRB protocol #001631001631) and all participants provided written informed consent to participate.

## Acknowledgements

This work used the Office of Cyber Infrastructure and Computational Biology High Performance Computing cluster at NIAID, Bethesda, MD. We thank Justin Lack for supporting bioinformatic analysis and Katherine Rees for critical reading of this manuscript.

