## Extended and Supplemental Figures for "Bismuth subsalicylate profoundly alters gut microbiome and immunity with increased susceptibility to infection"

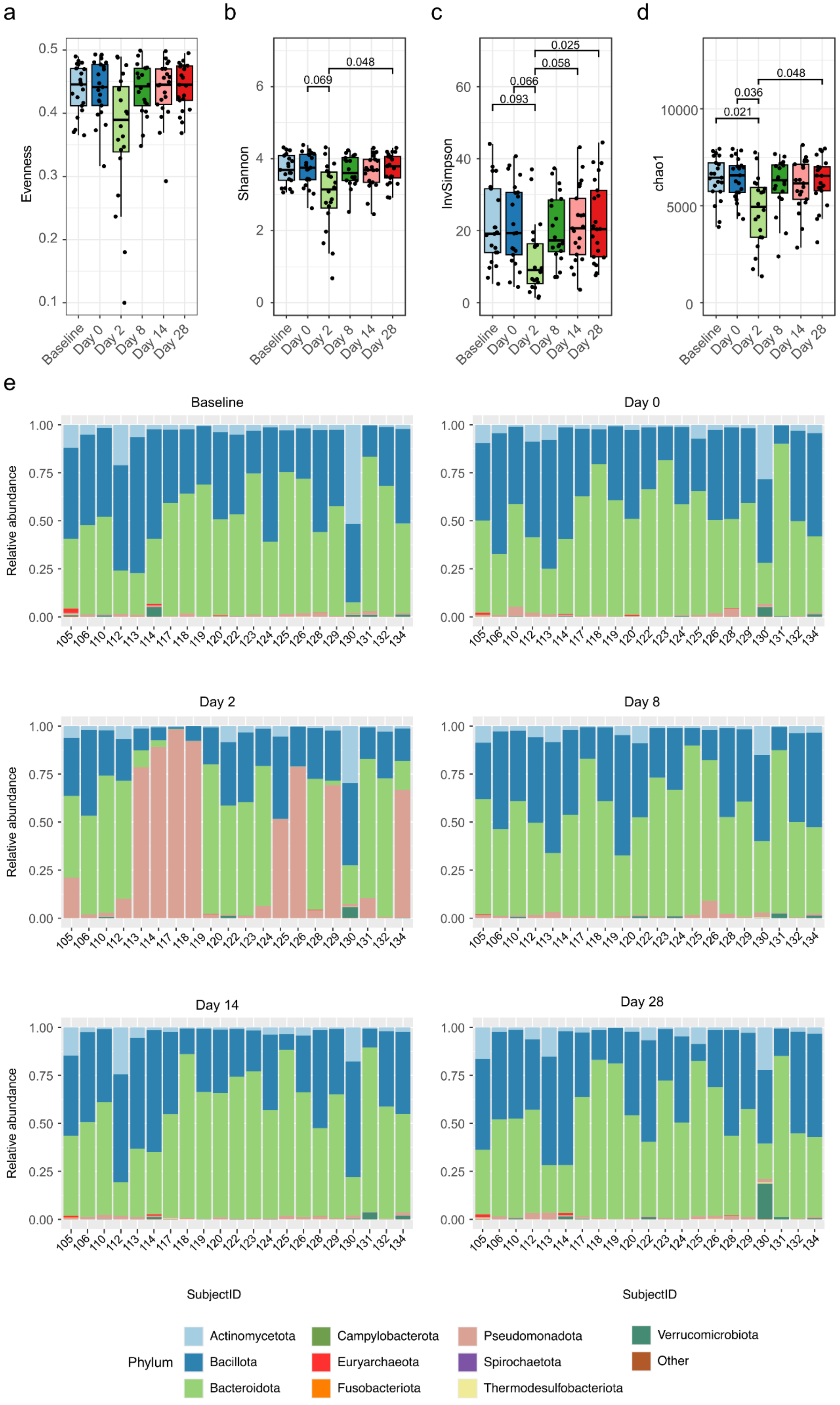

a
b
c
d
Evenness
Shannon
InvSimpson
chao1
0.069
0.048
0.093
0.066
0.058
0.025
0.021
0.036
0.048
Baseline
Day 0
Day 2
Day 8
Day 14
Day 28
e
Relative abundance
SubjectID
Phylum
Actinomycetota
Bacillota
Bacteroidota
Campylobacterota
Euryarchaeota
Fusobacteriota
Pseudomonadota
Spirochaetota
Thermodesulfobacteriota
Verrucomicrobiota
Other

**Extended Data Figure 1. Alpha Diversity metrics and phyla composition of fecal microbiome after BSS treatment. a-d**, Metrics of alpha diversity evenness (**a**), Shannon index (**b**), Inverse Simpson index (**c**) and chao1 index (**d**) at baseline and day 0, 2, 8, 14, 28. P values less than 0.1 shown, as calculated by Dunn's test. **e**, Relative abundance of top phyla at baseline and day 0, 2, 8, 14 and 28. Each individual subject is represented by subject identification number.

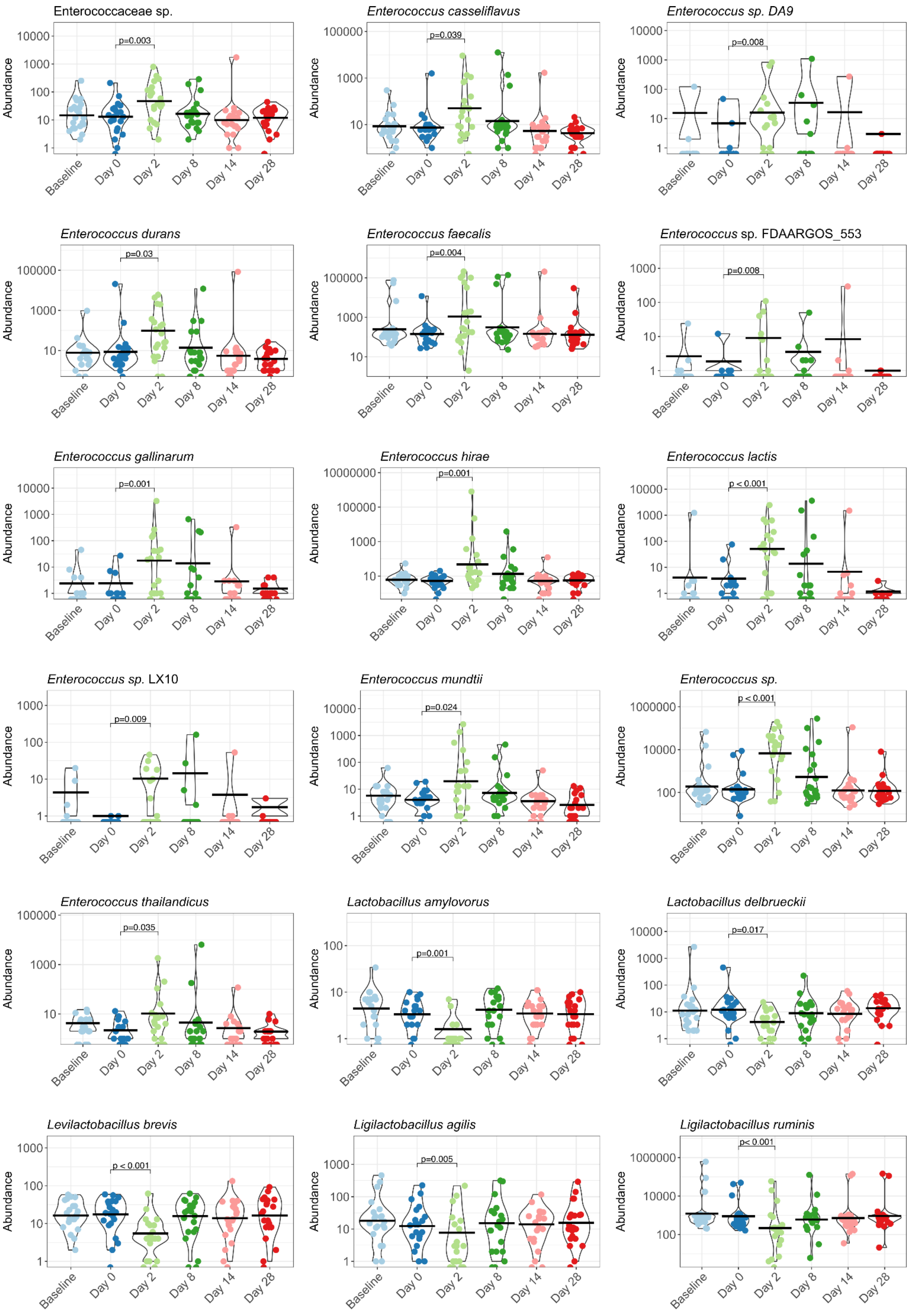
Enterococcaceae sp.
p=0.003
Enterococcus casseliflavus
p=0.039
Enterococcus sp. DA9
p=0.008
Enterococcus durans
p=0.03
Enterococcus faecalis
p=0.004
Enterococcus sp. FDAARGOS_553
p=0.008
Enterococcus gallinarum
p=0.001
Enterococcus hirae
p=0.001
Enterococcus lactis
p < 0.001
Enterococcus sp. LX10
p=0.009
Enterococcus mundtii
p=0.024
Enterococcus sp.
p < 0.001
Enterococcus thailandicus
p=0.035
Lactobacillus amylovorus
p=0.001
Lactobacillus delbrueckii
p=0.017
Levilactobacillus brevis
p < 0.001
Ligilactobacillus agilis
p=0.005
Ligilactobacillus ruminis
p< 0.001
Abundance
Baseline
Day 0
Day 2
Day 8
Day 14
Day 28

**Extended Data Figure 2. Relative Abundance of Enterococcus and Lactobacillus species**. Species of *Lactobacillus* and *Enterococcus* that are differentially abundant at day 2 compared to day 0 with a p value < 0.05, after adjustment for multiple comparisons with Benjamini-Hochberg procedure, and a coefficient (effect size) of < -1 or > 1.

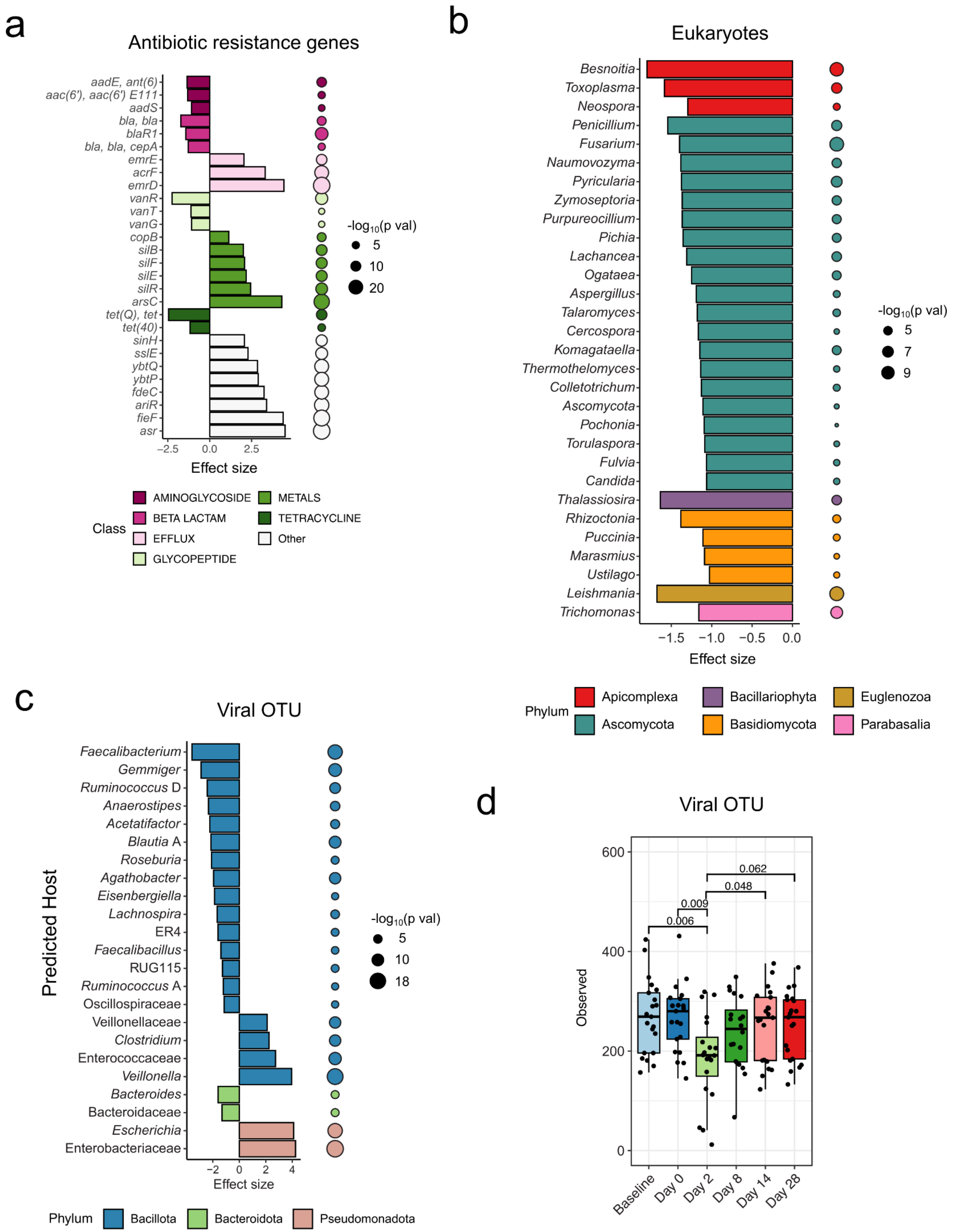
a
Antibiotic resistance genes
aadE, ant(6)
aac(6'), aac(6') E111
aadS
bla, bla
blaR1
bla, bla, cepA
emrE
acrF
emrD
vanR
vanT
vanG
copB
silB
silF
silE
silR
arsC
tet(Q), tet
tet(40)
sinH
sslE
ybtQ
ybtP
fdeC
ariR
fieF
asr
−2.5 0.0 2.5
Effect size
-log10(p val)
5
10
20
Class
AMINOGLYCOSIDE
BETA LACTAM
EFFLUX
GLYCOPEPTIDE
METALS
TETRACYCLINE
Other
b
Eukaryotes
Besnoitia
Toxoplasma
Neospora
Penicillium
Fusarium
Naumovozyma
Pyricularia
Zymoseptoria
Purpureocillium
Pichia
Lachancea
Ogataea
Aspergillus
Talaromyces
Cercospora
Komagataella
Thermothelomyces
Colletotrichum
Ascomycota
Pochonia
Torulaspora
Fulvia
Candida
Thalassiosira
Rhizoctonia
Puccinia
Marasmius
Ustilago
Leishmania
Trichomonas
−1.5 −1.0 −0.5 0.0
Effect size
-log10(p val)
5
7
9
Phylum
Apicomplexa
Ascomycota
Bacillariophyta
Basidiomycota
Euglenozoa
Parabasalia
c
Viral OTU
Predicted Host
Faecalibacterium
Gemmiger
Ruminococcus D
Anaerostipes
Acetatifactor
Blautia A
Roseburia
Agathobacter
Eisenbergiella
Lachnospira
ER4
Faecalibacillus
RUG115
Ruminococcus A
Oscillospiraceae
Veillonellaceae
Clostridium
Enterococcaceae
Veillonella
Bacteroides
Bacteroidaceae
Escherichia
Enterobacteriaceae
−2 0 2 4
Effect size
-log10(p val)
5
10
18
Phylum
Bacillota
Bacteroidota
Pseudomonadota
d
Viral OTU
Observed
600
400
200
0
0.062
0.048
0.009
0.006
Baseline
Day 0
Day 2
Day 8
Day 14
Day 28

**Extended Data Figure 3. Alterations in antibiotic resistance genes, eukaryotes and viruses after BSS treatment. a-c**, Differentially abundant antimicrobial resistance genes (**a**), eukaryotic taxa (**b**) and viral OTUs (**c**) at day 2 compared to day 0, with a p value $< 0.05$. P values were adjusted for multiple comparisons with Benjamini-Hochberg procedure, and a coefficient (effect size) of $< -1$ or $> 1$. Viral OTU in **c** represented by predicted bacterial host genus. Effect size represents coefficient produced from MaAslin2, representing strength and direction of relationship. **d**, Number of observed viral OTUs at each time point, with statistically significance differences shown with p values adjusted with Tukey's test for multiple comparisons.

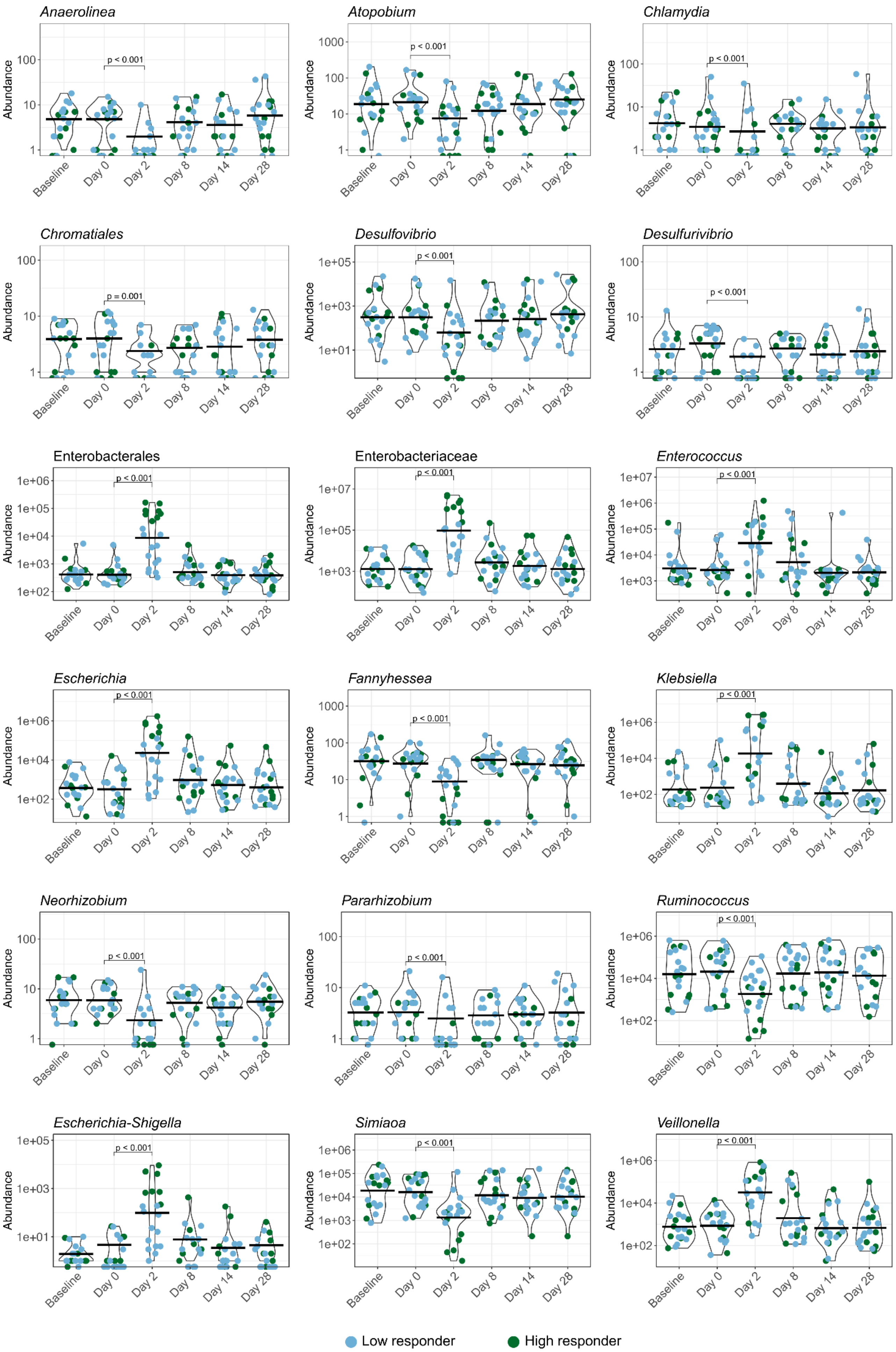

Anaerolinea
p < 0.001
Atopobium
p < 0.001
Chlamydia
p < 0.001
Chromatiales
p = 0.001
Desulfovibrio
p < 0.001
Desulfurivibrio
p < 0.001
Enterobacterales
p < 0.001
Enterobacteriaceae
p < 0.001
Enterococcus
p < 0.001
Escherichia
p < 0.001
Fannyhessea
p < 0.001
Klebsiella
p < 0.001
Neorhizobium
p < 0.001
Pararhizobium
p < 0.001
Ruminococcus
p < 0.001
Escherichia-Shigella
p < 0.001
Simiaoa
p < 0.001
Veillonella
p < 0.001
Abundance
Baseline
Day 0
Day 2
Day 8
Day 14
Day 28
Low responder
High responder

**Extended Data Figure 4. Select taxa altered in both high and low responders after BSS.** Abundance of genera that were differentially abundant in both high responders and low responders at day 2 compared to day 0 with a p value, adjusted for multiple comparisons with Benjamini-Hochberg procedure, of $< 0.20$ and coefficient (effect size) of $< -1.5$ or $> 1.5$.

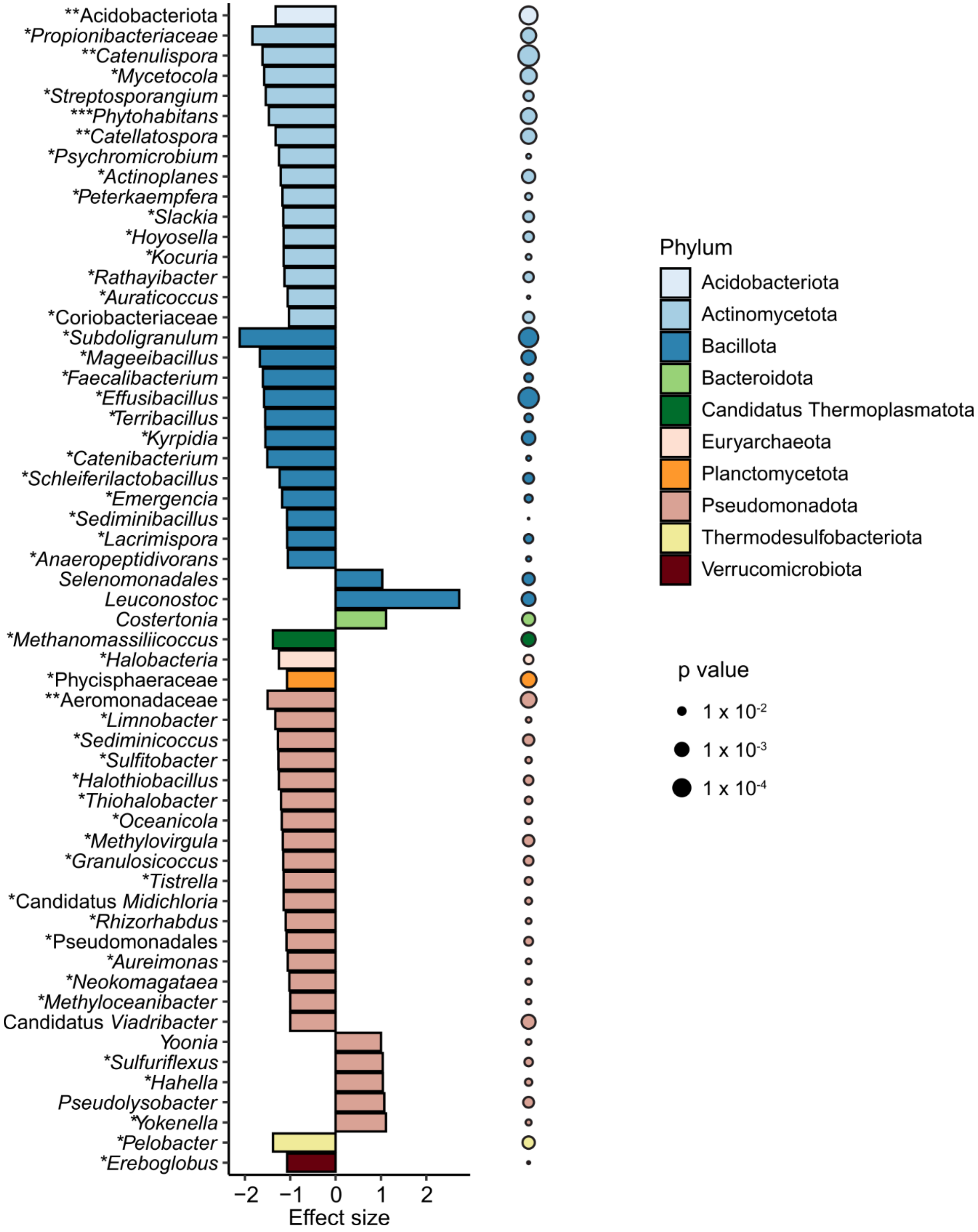


**Extended Data Figure 5. High responders showed lasting changes in microbiome composition.** Top differentially abundant genera at day 8 compared to day 0 within the subset of 'High responder' subjects. P values were adjusted for multiple comparisons with BH and differences with an effect size (coefficient from MaAslin2) < -1 or > 1 are shown. Effect size represents coefficient produced from MaAslin2, representing strength and direction of relationship Asterisk indicate taxa also passing significance threshold at day 2 (*), days 2 and 14 (**) and days 2,14 and 28 (***).

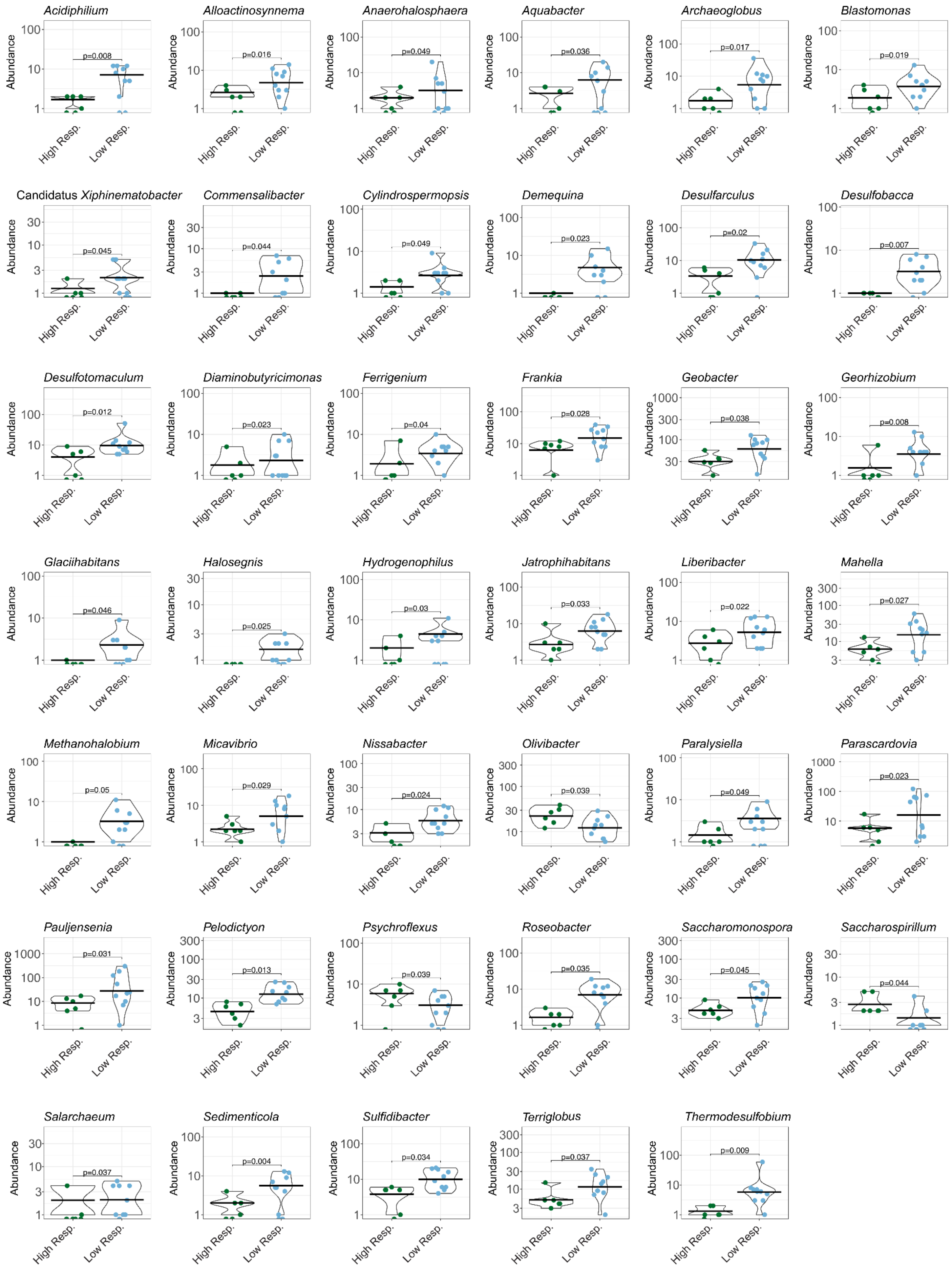

Acidiphilium p=0.008
Alloactinosynnema p=0.016
Anaerohalosphaera p=0.049
Aquabacter p=0.036
Archaeoglobus p=0.017
Blastomonas p=0.019
Candidatus Xiphinematobacter p=0.045
Commensalibacter p=0.044
Cylindrospermopsis p=0.049
Demequina p=0.023
Desulfarculus p=0.02
Desulfobacca p=0.007
Desulfotomaculum p=0.012
Diaminobutyricimonas p=0.023
Ferrigenium p=0.04
Frankia p=0.028
Geobacter p=0.038
Georhizobium p=0.008
Glaciihabitans p=0.046
Halosegnis p=0.025
Hydrogenophilus p=0.03
Jatrophihabitans p=0.033
Liberibacter p=0.022
Mahella p=0.027
Methanohalobium p=0.05
Micavibrio p=0.029
Nissabacter p=0.024
Olivibacter p=0.039
Paralysiella p=0.049
Parascardovia p=0.023
Pauljensenia p=0.031
Pelodictyon p=0.013
Psychroflexus p=0.039
Roseobacter p=0.035
Saccharomonospora p=0.045
Saccharospirillum p=0.044
Salarchaeum p=0.037
Sedimenticola p=0.004
Sulfidibacter p=0.034
Terriglobus p=0.037
Thermodesulfobium p=0.009
Abundance
High Resp.
Low Resp.

**Extended Data Figure 6. Fecal microbiome taxa at day 0 that predict responder status to BSS.** Abundances of genera at day 0 that are predictive of a high or low response to BSS observed at day 2, as calculated by ROC analysis passing a p value threshold of <0.05. Reported p values are from t tests on fold change differences between high and low responders.

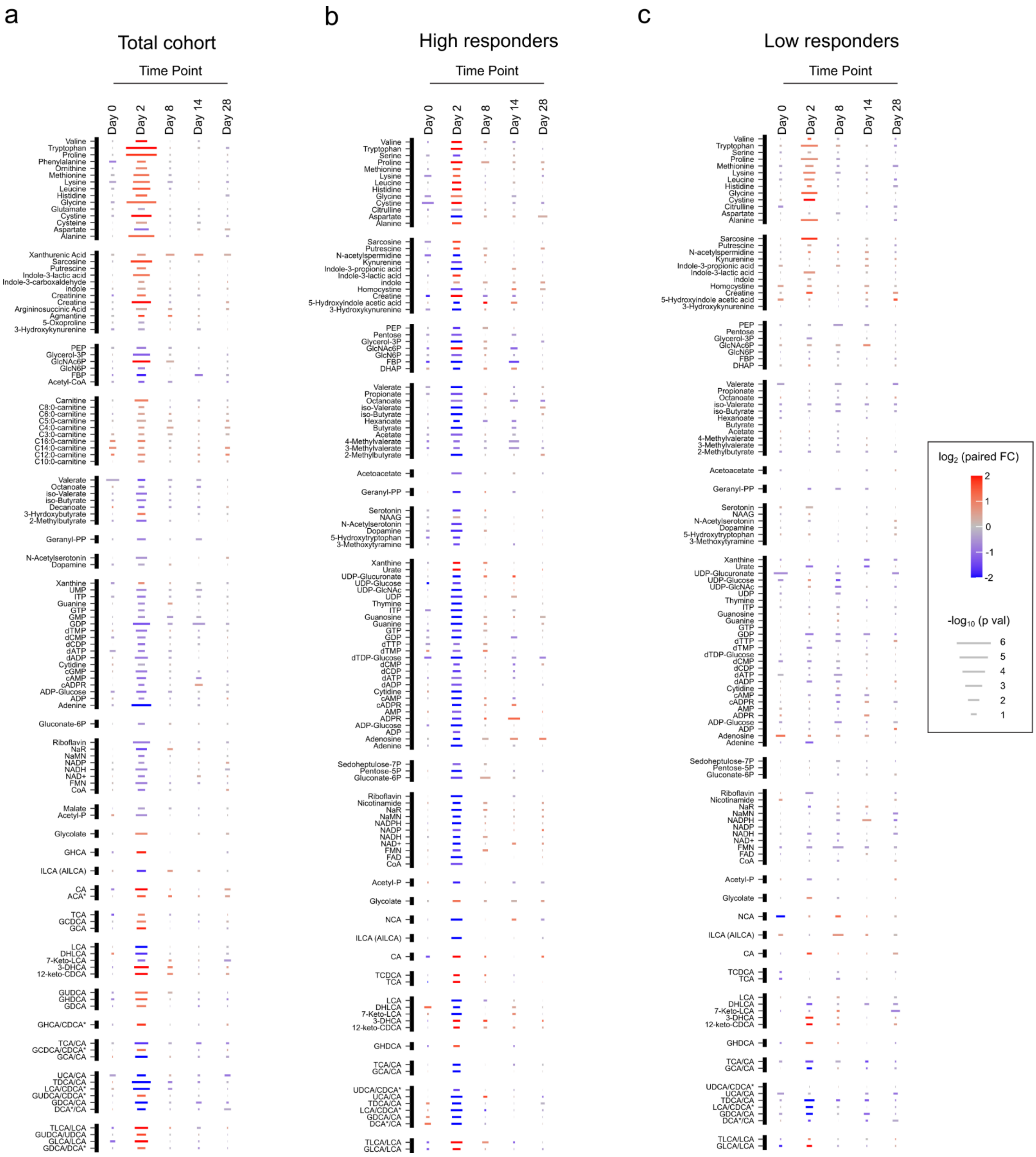

a
Total cohort
Time Point
Day 0
Day 2
Day 8
Day 14
Day 28
Valine
Tryptophan
Proline
Phenylalanine
Ornithine
Methionine
Lysine
Leucine
Histidine
Glycine
Glutamate
Cystine
Cysteine
Aspartate
Alanine
Xanthurenic Acid
Sarcosine
Putrescine
Indole-3-lactic acid
Indole-3-carboxaldehyde
indole
Creatinine
Creatine
Argininosuccinic Acid
Agmatine
5-Oxoproline
3-Hydroxykynurenine
PEP
Glycerol-3P
GlcNAc6P
GlcN6P
FBP
Acetyl-CoA
Carnitine
C8:0-carnitine
C6:0-carnitine
C5:0-carnitine
C4:0-carnitine
C3:0-carnitine
C16:0-carnitine
C14:0-carnitine
C12:0-carnitine
C10:0-carnitine
Valerate
Octanoate
iso-Valerate
iso-Butyrate
Decanoate
3-Hyrdoxybutyrate
2-Methylbutyrate
Geranyl-PP
N-Acetylserotonin
Dopamine
Xanthine
UMP
ITP
Guanine
GTP
GMP
GDP
dTMP
dCMP
dCDP
dATP
dADP
Cytidine
cGMP
cAMP
cADPR
ADP-Glucose
ADP
Adenine
Gluconate-6P
Riboflavin
NaR
NaMN
NADP
NADH
NAD+
FMN
CoA
Malate
Acetyl-P
Glycolate
GHCA
ILCA (AILCA)
CA
ACA*
TCA
GCDCA
GCA
LCA
DHLCA
7-Keto-LCA
3-DHCA
12-keto-CDCA
GUDCA
GHDCA
GDCA
GHCA/CDCA*
TCA/CA
GCDCA/CDCA*
GCA/CA
UCA/CA
TDCA/CA
LCA/CDCA*
GUDCA/CDCA*
GDCA/CA
DCA*/CA
TLCA/LCA
GUDCA/UDCA
GLCA/LCA
GDCA/DCA*
b
High responders
Time Point
Day 0
Day 2
Day 8
Day 14
Day 28
Valine
Tryptophan
Serine
Proline
Methionine
Lysine
Leucine
Histidine
Glycine
Cystine
Citrulline
Aspartate
Alanine
Sarcosine
Putrescine
N-acetylspermidine
Kynurenine
Indole-3-propionic acid
Indole-3-lactic acid
indole
Homocysteine
Creatine
5-Hydroxyindole acetic acid
3-Hydroxykynurenine
PEP
Pentose
Glycerol-3P
GlcNAc6P
GlcN6P
FBP
DHAP
Valerate
Propionate
Octanoate
iso-Valerate
iso-Butyrate
Hexanoate
Butyrate
Acetate
4-Methylvalerate
3-Methylvalerate
2-Methylbutyrate
Acetoacetate
Geranyl-PP
Serotonin
NAAG
N-Acetylserotonin
Dopamine
5-Hydroxytryptophan
3-Methoxytyramine
Xanthine
Urate
UDP-Glucuronate
UDP-Glucose
UDP-GlcNAc
UDP
Thymine
ITP
Guanosine
Guanine
GTP
GDP
dTTP
dTMP
dTDP-Glucose
dCMP
dCDP
dATP
dADP
Cytidine
cAMP
cADPR
AMP
ADPR
ADP-Glucose
ADP
Adenosine
Adenine
Sedoheptulose-7P
Pentose-5P
Gluconate-6P
Riboflavin
Nicotinamide
NaR
NaMN
NADPH
NADP
NADH
NAD+
FMN
FAD
CoA
Acetyl-P
Glycolate
NCA
ILCA (AILCA)
CA
TCDCA
TCA
LCA
DHLCA
7-Keto-LCA
3-DHCA
12-keto-CDCA
GHDCA
TCA/CA
GCA/CA
UDCA/CDCA*
UCA/CA
TDCA/CA
LCA/CDCA*
GDCA/CA
DCA*/CA
TLCA/LCA
GLCA/LCA
c
Low responders
Time Point
Day 0
Day 2
Day 8
Day 14
Day 28
Valine
Tryptophan
Serine
Proline
Methionine
Lysine
Leucine
Histidine
Glycine
Cystine
Citrulline
Aspartate
Alanine
Sarcosine
Putrescine
N-acetylspermidine
Kynurenine
Indole-3-propionic acid
Indole-3-lactic acid
indole
Homocysteine
Creatine
5-Hydroxyindole acetic acid
3-Hydroxykynurenine
PEP
Pentose
Glycerol-3P
GlcNAc6P
GlcN6P
FBP
DHAP
Valerate
Propionate
Octanoate
iso-Valerate
iso-Butyrate
Hexanoate
Butyrate
Acetate
4-Methylvalerate
3-Methylvalerate
2-Methylbutyrate
Acetoacetate
Geranyl-PP
Serotonin
NAAG
N-Acetylserotonin
Dopamine
5-Hydroxytryptophan
3-Methoxytyramine
Xanthine
Urate
UDP-Glucuronate
UDP-Glucose
UDP-GlcNAc
UDP
Thymine
ITP
Guanosine
Guanine
GTP
GDP
dTTP
dTMP
dTDP-Glucose
dCMP
dCDP
dATP
dADP
Cytidine
cAMP
cADPR
AMP
ADPR
ADP-Glucose
ADP
Adenosine
Adenine
Sedoheptulose-7P
Pentose-5P
Gluconate-6P
Riboflavin
Nicotinamide
NaR
NaMN
NADPH
NADP
NADH
NAD+
FMN
FAD
CoA
Acetyl-P
Glycolate
NCA
ILCA (AILCA)
CA
TCDCA
TCA
LCA
DHLCA
7-Keto-LCA
3-DHCA
12-keto-CDCA
GHDCA
TCA/CA
GCA/CA
UDCA/CDCA*
UCA/CA
TDCA/CA
LCA/CDCA*
GDCA/CA
DCA*/CA
TLCA/LCA
GLCA/LCA
log2 (paired FC)
2
1
0
-1
-2
-log10 (p val)
6
5
4
3
2
1

**Extended Data Figure 7. Fecal metabolic changes in high and low responder patients. a-c,** Line plot of significantly altered metabolites grouped by corresponding pathway, in all subjects (**a**) only high responders (**b**) or only low responders (**c**). Fold changes were calculated using paired time point samples, Day2/Day0. Line color reflected the log2 transformed fold changes. Significant difference between paired time point samples were calculated using the Wilcoxon test. Metabolites included in the plot passed an FDR 10% significance filter after adjustment using the Benjamini-Hochberg method. Line length reflected the -log10 transformed adjusted p-values.

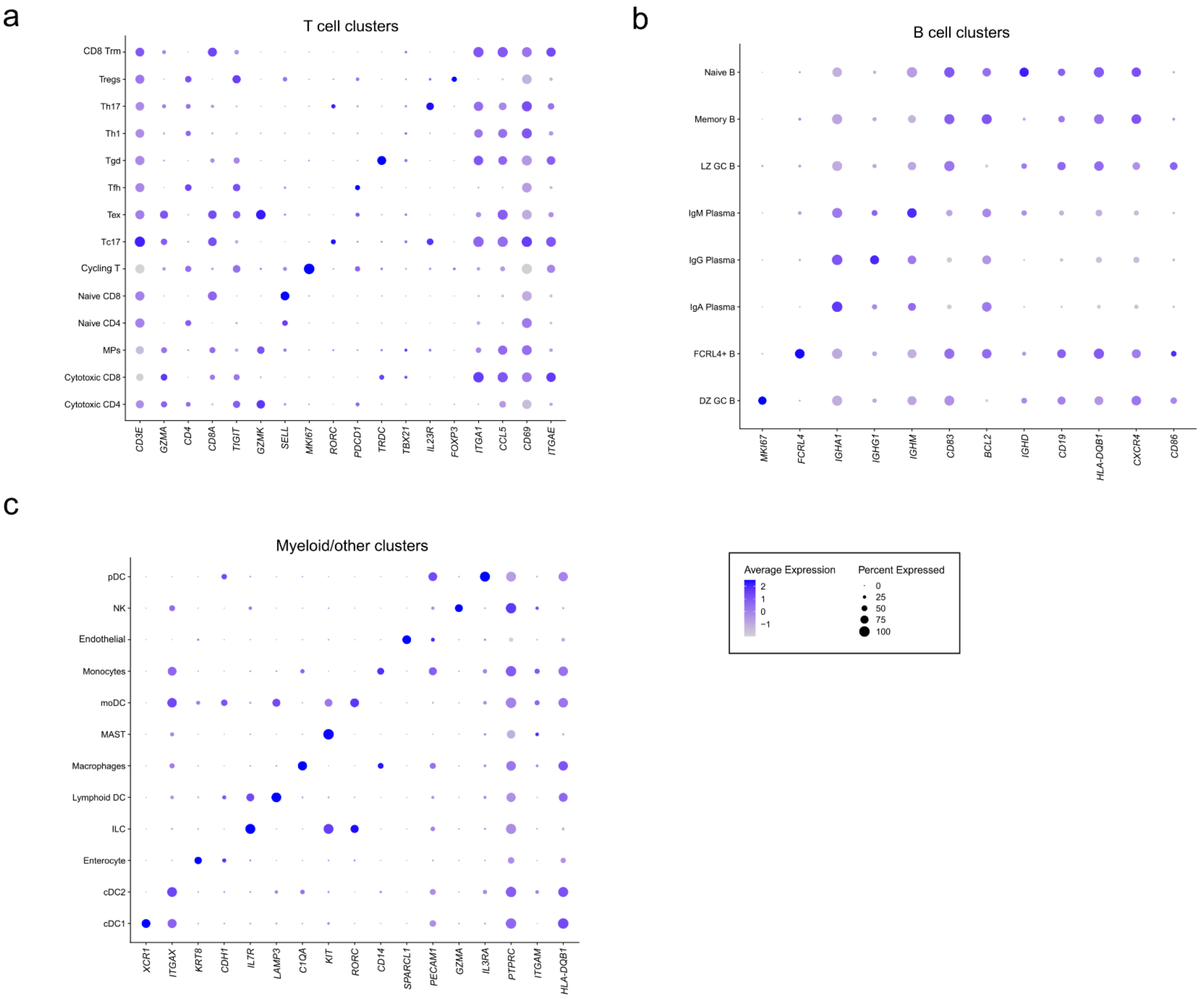


**Extended Data Figure 8. Select gene expression markers for each identified cluster in ileal scRNAseq. a-c,** Bubble plots of each cell subset of (**a**) T cells, (**b**) B cells, and (**c**) myeloid and other cell subsets. Chosen genes representative of gene sets used to identify each cluster. Color of circles represents average expression and size of circles represents percent expressing each gene.

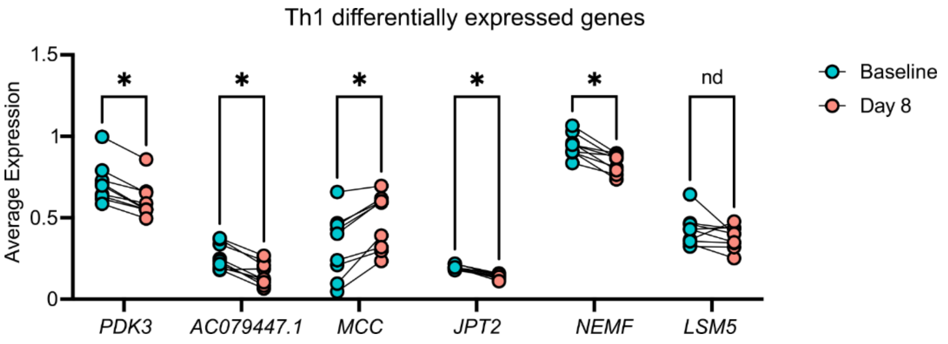


**Extended Data Figure 9. DEGs for Th1 cluster in ileal scRNAseq.** Significantly differentially expressed genes in the Th1 cluster at day 8 of BSS treatment, as calculated by DESeq2 pseudobulk analysis. Genes were also compared by paired t test with FDR correction, * indicates genes which passed FDR threshold, nd indicates genes which did not pass FDR threshold.

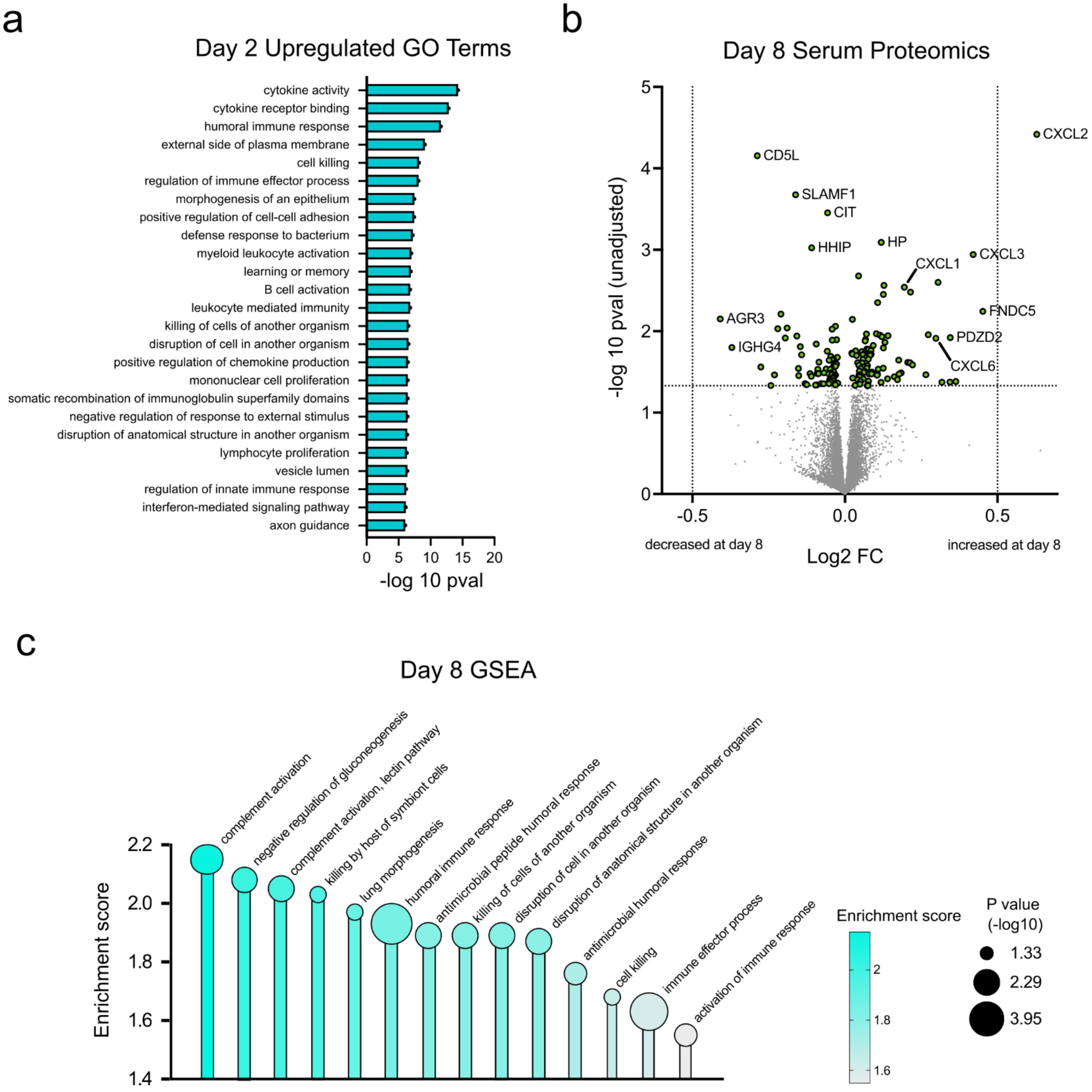


**Extended Data Figure 10. Alterations in serum proteome at days 2 and 8 after BSS treatment. a**, differentially upregulated GO terms from serum proteomics at day 2 of BSS treatment compared to day 0. **b**, volcano plot of serum proteins at day 8 post BSS treatment compared to day 0. No proteins passed FDR threshold, p values shown are not adjusted for multiple comparisons. **c**, Gene set enrichment analysis of day 8 serum proteins compared to day 0, with all significantly upregulated pathways shown at day 8 (no pathways were significantly downregulated).

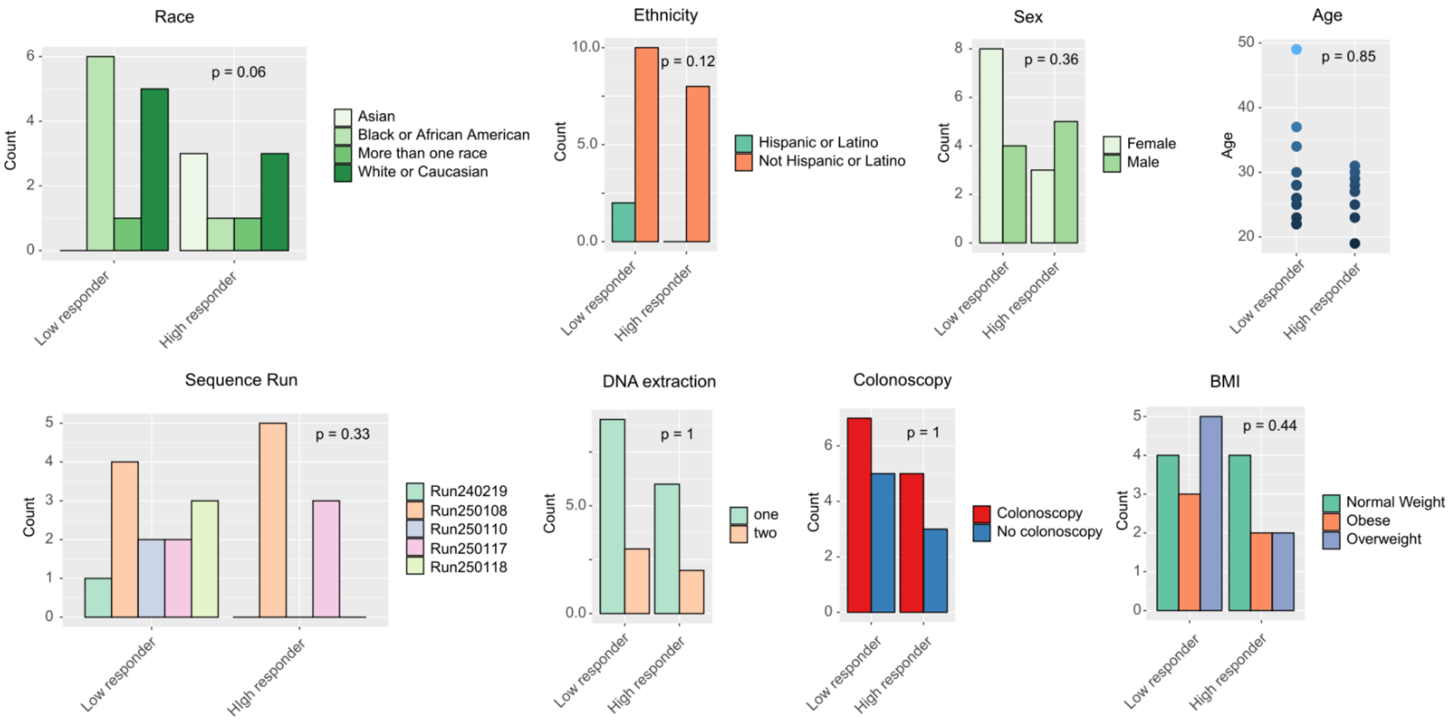


**Supplementary Figure 1. No correlations between patient metadata and microbiome response to BSS.** Collected patient metadata was correlated to responder status as defined by beta diversity of the microbiome and Pseudomonadota frequency in Fig. 3a. Displayed p values were calculated with Fisher's exact test and none passed significance threshold of $p < 0.05$.

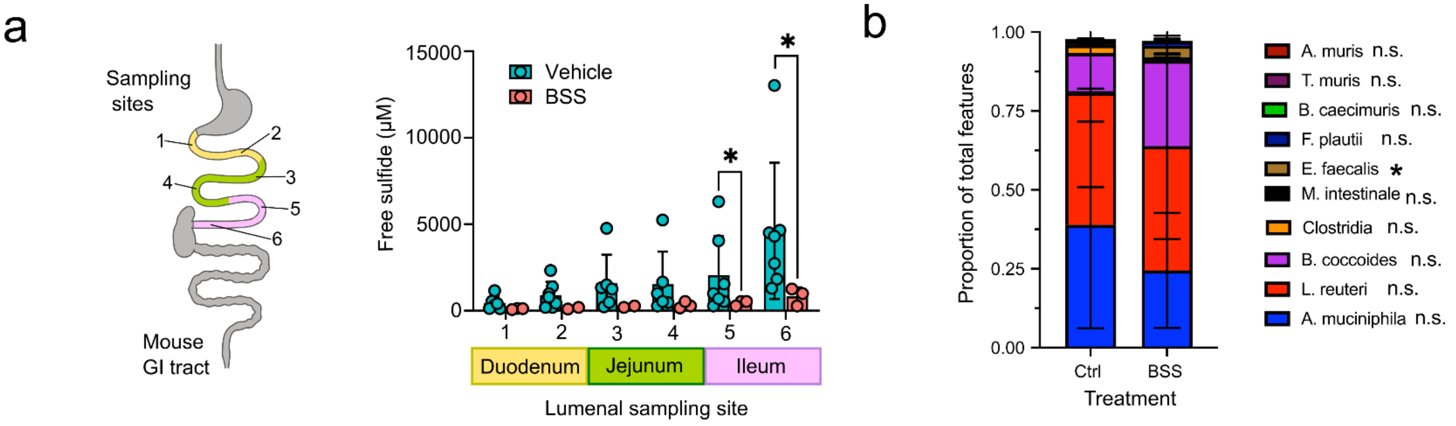


**Supplementary Figure 2. Sulfide concentration and compositional change in BSS treated OMM12 gnotobiotic mice. a**, Free hydrogen sulfide concentration in the lumen of wild type SPF mouse small intestine sampled at various sites and measured by microelectrode. Mice were treated with vehicle or BSS by gavage for 3 days before measurements. **b,** Composition of fecal microbiome in OMM12-colonized gnotobiotic mice 3 days after BSS or vehicle treatment. * $p < 0.05$, n.s. not significant, unpaired t test.
